# Not all women are equally at risk: A Demographic health survey (DHS) 2023 based analysis of overweight and obesity inequalities among women in the Democratic Republic of the Congo

**DOI:** 10.64898/2026.08.19.26360799

**Authors:** Baperman Abdel Aziz Siri, Jean Shonganye, Papy Musas, Bien Aime Mandja, George Mutuale, Joseph B. Otshudiandjeka, Dieudonné Mwanba Kazadi

**Author notes:** Corresponding author: Siri Baperman Abdel Aziz.

## Abstract

**Background:** In sub-Saharan Africa, women are navigating overlapping burdens of undernutrition and rising overweight/obesity, often within fragile health system and rapidly changing food environments. In the DRC, theses tensions may be intensified by rapid urbanization, socioeconomic disparities, insecurity and shifting lifestyles. Despite those changes, national level evidence on who is the most affected by excess weight and why remains scarce. This study assessed the determinant of overweight and obesity among Congolese women of reproductive age, aiming to highlight the social and geographic inequalities.

**Methods:** We analysed nationally representative data from the 2023 DHS. Thes study included 10,740 non-pregnant women aged 15-49 years with valid anthropometry. Overweight/obesity was defined as BMI ≥ 25 Kg/m2. We examined a wide broad range of potential associated factors, including province, residence, socioeconomic status, household structure, education level, marital status, occupation, dietary diversity score, healthy diet related indicators, media exposure, internet use and health service utilisation. Weighted analyses accounted for the DHS sampling design. Variables associated at p value < 0.20 were retained for multivariable modelling. Multicollinearity was assed via adjusted GVIFs. Four hierarchical weighted logistic regression were built; the fully adjusted model guided final interpretation.

**Results:** Nearly on five women of reproductive age (19.5%) lived overweight or obesity. However, this burden was not evenly distributed. Women from Kongo Central and Tshuapa exhibited significantly lower odds, while those in Bas-Uele, Nord-Kivu, Sud-Kivu and Maniema were substantially more affected, highlighting spatial inequities. Women living in rural areas had lower odds of overweight/obesity compared with their urban counterparts (aOR=0.6; 95% CI: 0.48-0.79; p<0.001).

A pronounced socioecomic gradient was observed. Compared with the poorest households, the likelihood of excess weight increases progressively among women in middle income household (aOR=1.65;95% CI:1.13-2.41), rich households (aOR=2.41; 95%CI:1.62-3.60), and was highest among the richest (aOR=4.19; 95%CI: 2.45-7.16). Larger households appeared protective, with lower odds observed in household of 4-5 members (aOR=0.68; 95%CI:0.5-0.92), 6-7 (aOR=0.72;95% CI: 0.54-0.97) and ≥8 members (aOR=0.69; 95%CI:0.50-0.95) compared with smaller household.

Age was the strongest predictor, with risk sharply accelerating after 30 years. Being married or in union was associated with higher odds. Notably, frequent internet use independently predicted overweight/obesity. In contrast, dietary diversity and unhealthy food indicators were not significantly significant in the fully adjusted models.

**Conclusion:** Overweight and obesity are rising among Congolese women, but unevenly and unjustly. Urban residence, socioeconomic status, age and digital exposure strongly sharply shape who is the most affected, revealing deep social and geographic inequities. Addressing this growing epidemic requires equity-oriented, province specific actions, alongside stronger primary prevention.

## 1. Background

Overweight and obesity have become major public health challenges of the 21st century. Globally, the number of adults affected has more than doubled since 1990, reaching estimated 2.6 billion in 2022, including over 890 million living with obesity (1,2). Although historically concentrated in high income countries, excess body weight is now increasing most rapidly in low and middle income countries, driven by demographic change, accelerated urbanisation, and profound transformation in food systems and lifestyles (3,4,5,6).

Sub-Saharan Africa is increasingly at the centre of this transition. The coexistence of persistent undernutrition and rising overweight and obesity has given rise to a double burden of malnutrition, posing complex challenges for health systems that remain largely oriented toward deficiency-related conditions (7,8,9). Women of reproductive age are particularly affected, due to biological, social, and structural vulnerabilities operating across the life course (10,11).

Evidence from DHS and population-based studies consistently demonstrates that overweight and obesity in Sub Saharan are socially patterned. Women, urban residents, and individuals from wealthier household bear a disproportionate share of the burden (12,13,14,15,16).

Across multiple countries, age, household wealth, place of residence, marital status, and occupation emerge as robust correlates of excess body weight, underscoring the central role of contextual and structural determinants rather than individual behaviours alone (13,17,18,19). Cross-country analyses further indicate that many African countries are in a early stage of the of nutrition and obesity transition characterised by a monotonic increase in obesity risk across wealth quintiles (6, 20). This pattern contrasts with later stages observed elsewhere, where the burden progressively shifts toward poorer populations, highlighting the dynamic and evolving nature of excess weight related inequalities in the region (3,20).

In the Democratic Republic of the Congo (DRC), signs of this nutritional transition are increasingly evident but remain insufficiently characterised. The 2013-2014 DHS reported that 15% of women in reproductive age living in urban areas were overweight or obese (21). More recent findings from the 2023 DHS indicate a further increase, with approximatively one five women aged 20-49 years (22.9%) affected, and substantially higher prevalence in urban settings (22). Prior local studies also point to marked heterogeneity by age, sex, and place of residence with high burdens among adult women in urban and peri urban context (23,24).

Despite this rising burden, national evidence on the determinants of overweight and obesity among Congolese women remains limited. Existing DHSs are largely descriptive, and any study have examined how contextual, household, sociodemographic, and behavioural factors jointly influence excess body weight using nationally representative data. This gap is critical, as evidence from other African settings suggests that early identification of these determinants is essential to prevent excess weight from becoming entrenched among more socioeconomically vulnerable groups (3,20,25).

This study estimates the prevalence of overweight and obesity among women of reproductive age in the Democratic republic of Congo and examines associated contextual, household, individual, and behavioural factors using nationally representative data from the 2023 DHS through multivariable analyses informed socio-ecological framework(26).

## 2. Method

### Study design, data source

We conducted a cross-sectional analytical study using data from the 2023 Demographic and Health Survey (DHS) of the Democratic Republic of Congo, which employs a stratified, two stage cluster sampling design to ensure national representativeness. We used individual recode (IR). All measurement and data collect followed DHS standard procedures(22).

### Study population

The target population included women aged 15-49 years. Inclusion criteria are women aged 15-49 years with valid measurements for weight and height (allowing BMI computation).

We excluded pregnant women and those with six months postpartum. We also excluded respondents with missing or implausible anthropometry. Body mass index with values < 16 or > 60 Kg/m2 were set to missing.

### Sampling

The 2023 DRC DHS followed the standard DHS protocol. In the first stage, enumeration areas were selected as primary sampling units within strata defined by province and urban-rural residence. In the second stage, households were systematically sampled within each cluster. All eligible women aged 15-49 years who were usual residents or present the night before the survey were invited to participate. A subsample was selected for anthropometry. The national sample included 26,520 households from 780 clusters with around 27100 women successfully interviewed(22).

### Study variables

The outcome was overweight/obesity, defined as A binary indicator was created, coded 1 for BMI ≥ 25 Kg/m^2^. and 0 otherwise.

#### Explanatory variables

Variables were selected based on previous studies and grouped conceptually (2,5,7,10,20,21,27,28).

#### Contextual factors

Province was recoded as categorical. Residence was coded as urban versus rural. The household index was categorised into five orders DHS quintiles: poorest, poor, middle, rich, and richest.

#### Household level variables

Household characteristics were derived from IR but correspond to the household questionnaire variable. Electricity availability was converted to categorial factors (0=No, 1=Yes). Household size was regrouped into four categories (1-3. 4-5, 6-7, ≥ 8 persons). Age of the household head was categorized into < 30, 30-39, 40-49, and ≥ 50 years. Sex of the household head was recoded as male versus female.

#### Individual level variables

Individual sociodemographic variables included age group categorised into five groups: 15-19, 20-24, 25-29, 30-34, 35-39, 40-44 and 45-49 years.

Although WHO recommends BMI for age (29) to assess nutritional status among adolescents, DHS reporting and much of the DHS based literature commonly apply adult BMI cut-offs to women 15-49 years to maintain comparability with DHS reporting and programmatic indictors. To assess robustness and the potential influence of age composition, we conducted analysis after restricting the sample to women aged 20-49 years, and we estimated overweight/obesity prevalence separately among adolescents aged 15-19 years.

Educational attainment was categorised as: no education, primary, secondary or tertiary. Marital status was classified into never married, married, in union, widowed, divorced, or separated.

Current work status was coded as working versus not working. Occupation was grouped into categories including professional, clerical, sales, agriculture, domestic, services, skilled manual and unskilled manual. Number of living children was derived into 0, 1-2,3-4, and ≥ 5 children. Additional variables include frequency of media exposure (internet, television, radio), categorised as never, less than once per week, at least once per week and almost every day. Health insurance coverage as well contraceptive use was coded as binary variables (yes/no).

#### Nutrition and health practice

A minimum dietary diversity score (DDS) was assessed using DHS questions on food group consumption. ten mutually exclusive food groups were constructed according to the minimum diversity for women : 1) grain, white roots and tubers; 2) pulses; 3)nuts and seeds; 4)dairy ;5) meat, poultry and fish ; 6) eggs; 7) dark green leafy vegetables; 8) Vitamin A rich fruits and vegetables ; 9) other vegetables; and 10) other fruits. A dietary diversity score ranging from 0 to 10 was generated, and a binary variable indicating adequate diversity was defined as consumption of at least five food groups.

An unhealthy food score was constructed based on reported consumption of sugar foods, fried foods, sugar sweetened beverages, and processed meats. Each component was coded 1 if consumed and o otherwise and summed to obtain an unhealthy score (0–4). A binary indicator (unhealthy any) captured any consumption of unhealthy food (score > 0).

Health care seeking in the last 12 months was recoded as binary variable (yes/no).

### Statistical analysis

Survey design was specified using R survey package, with primary sampling units, stratification, and sampling weight defined in accordance with DHS guidelines. All analyses accounted for clustering, stratification, and weights. For continuous variable, we computed survey weighted medians and interquartile ranges (Q1, Q3).

#### Descriptive analysis

We first described the distribution of all explanatory variables in the overall sample. For categorical variables, weighted proportions and 95% confidence intervals were estimated. For continuous variables

#### Bivariate analysis

Each explanatory variable was examined in bivariate survey weighted regression with overweight/obesity as the dependent variable, using svyglm with a quasibinomial family. For each variable, crude odds ratio with 95% CI were assessed using survey weighted logistic regression. Variables with p < 0.20 (Wald test) were retained as candidates for the multivariable modelling after checking for collinearity. A threshold of generalized variance inflation factors (GVIF) adjusted ≥ 2.2 was used to indicate problematic collinearity.

#### Multivariate hierarchical analysis

A series of four nested survey weighted logistic regression models with overweight/obesity as the outcome were built sequentially.

Model I (contextual): province, residence.

Model II (Household): addition of household characteristics.

Model III (individual): addition of sociodemographic factors.

Model IV (Nutrition/health): addition of nutritional and health related variables.

Let Yi denote the binary indicator for overweight or obesity for woman i. The final survey weighted logistic regression model was specified as follows:

*Logit [P(Yi=1)] = β0 + βc^T^ Xci + βh^T^ Xhi + βi^T^ Xii + βn^T^ Xni*, where *Xci, Xhi,Xii and Xni* represent contextual, individual and nutrition or health related covariates, respectively. All categorical variables were modelled using indicator (dummy) variables, with reference categories defined a priori reported in table 2. Adjusted odds ratio was obtained by exponentiating the regression coefficients (aOR=e^β^).

Model improvement was assessment using McFadden’s pseudo R^2^, deviance, and the change in pseudo R^2^(ΔR²). The model showing the largest improvement was retained as the final model. Adjusted odds ratio (aOR) with 95% confidence intervals were reported.

### Ethical considerations

**Figure 1:**
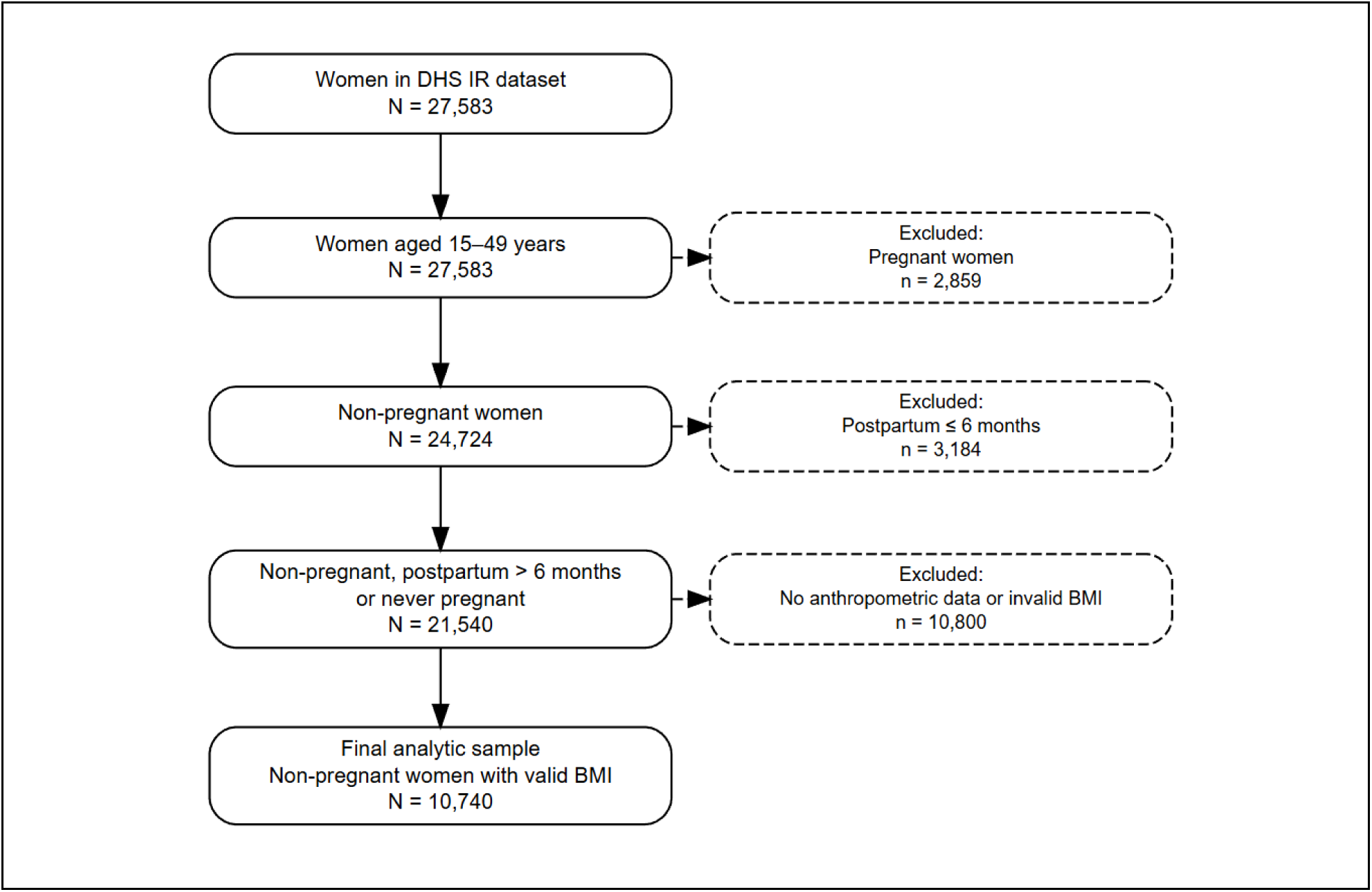
Flow diagram of the study sample selection from the 2023 DRC Demographic and health survey.

This study is a secondary analysis of anonymized DHS data. We obtained permission to use and analyse the DHS data form the DHS program (ICF) (Ref: 214022). The original survey received ethical approval from national authorities and ICF’s Institutional review board. Informed consent was obtained from all participants during primary data collection. No personal identifiers were available in the dataset. The present study complies with the principles of the Declaration of Helsinki and ensures confidentiality throughout.

## Results

### Background of study participants

A total of 10740 women aged 15–49-yearswere included in the study. They came from the 26 regions of the country, with 15.9% living in Kinshasa and 8.8% in Nord Kivu. More than half of the women resided in rural areas (56.2%). Household wealth varied: one in four lived in richest households (25.7%), while about one in six were in the poorest or poorer categories. Most women lived in homes without electricity (73.3%). The average age was 28.5 years. Over half had completed secondary school (56%), and two in five had (40%) had never been married. Nearly two out of five women had no children, and almost half were not working at the time of the survey.

Patterns of information access and health coverage highlight that the majority did not use internet (83%), did not listen to the radio (67.8%), and did not watch television (66.9%). Most were not covered by any form of health insurance (95.6%) and were not using contraceptive methods (82,9%). Dietary indicators revealed that many had inadequate dietary diversity, while about one in three reports consuming unhealthy foods (34.6%) (Table 1).

**Table 1:** Characteristics of the study population.

| Variable | n<br>unweighted | N<br>Weighted | Unweighted<br>(%) | Weighted %<br>(IC%95) |
| --- | --- | --- | --- | --- |
| **Province** |  |  |  |  |
| Kinshasa | 619 | 10740 | 5.8 | 15.9 [13.4- 18.4] |
| Kongo Central | 441 | 10740 | 4.1 | 6.9 [4.8- 9.1] |
| Kwango | 325 | 10740 | 3.0 | 2.4 [1.8- 3.0] |
| Kwilu | 454 | 10740 | 4.2 | 5.8 [3.9- 7.7] |
| Mai-Ndombe | 337 | 10740 | 3.1 | 1.7 [1.0- 2.5] |
| Equateur | 405 | 10740 | 3.8 | 3.2 [2.0- 4.5] |
| Nord Ubangi | 362 | 10740 | 3.4 | 1.3 [0.7- 1.9] |
| Sud Ubangi | 517 | 10740 | 4.8 | 2.6 [1.6- 3.7] |
| Mongala | 365 | 10740 | 3.4 | 1.6 [0.9- 2.3] |
| Tshuapa | 276 | 10740 | 2.6 | 1.4 [0.7- 2.0] |
| Tshopo | 362 | 10740 | 3.4 | 1.6 [0.8- 2.4] |
| Bas-Uele | 282 | 10740 | 2.6 | 1.0 [0.6- 1.4] |
| Haut Uele | 255 | 10740 | 2.4 | 1.8 [0.5- 3.1] |
| Ituri | 542 | 10740 | 5.0 | 5.0 [3.3- 6.8] |
| Nord-Kivu | 563 | 10740 | 5.2 | 8.8 [5.5- 12.1] |
| Sud-Kivu | 537 | 10740 | 5.0 | 7.2 [3.6- 10.9] |
| Maniema | 379 | 10740 | 3.5 | 2.7 [1.9- 3.6] |
| Haut-Katanga | 479 | 10740 | 4.5 | 6.4 [3.6- 9.2] |
| Haut Lomami | 472 | 10740 | 4.4 | 2.7 [1.8- 3.6] |
| Lualaba | 372 | 10740 | 3.5 | 3.3 [1.7- 5.0] |
| Tanganyika | 406 | 10740 | 3.8 | 2.4 [1.3- 3.5] |
| Lomami | 390 | 10740 | 3.6 | 2.5 [1.8- 3.1] |
| Sankuru | 286 | 10740 | 2.7 | 1.3 [0.8- 1.8] |
| Kasaï Oriental | 391 | 10740 | 3.6 | 3.2 [2.0- 4.4] |
| Kasaï | 446 | 10740 | 4.2 | 3.8 [2.0- 5.5] |
| Kasaï Central | 477 | 10740 | 4.4 | 3.4 [2.2- 4.6] |
| **Residence area** |  |  |  |  |
| Urban | 3817 | 10740 | 35.5 | 43.8 [39.4- 48.1] |
| Rural | 6923 | 10740 | 64.5 | 56.2 [51.9- 60.6] |
| **Wealth index** |  |  |  |  |
| Poorest | 2499 | 10740 | 23.3 | 16.5 [14.5- 18.5] |
| Poorer | 2417 | 10740 | 22.5 | 17.0 [15.2- 18.9] |
| Middle | 2341 | 10740 | 21.8 | 19.0 [16.8- 21.3] |
| Richer | 1970 | 10740 | 18.3 | 21.7 [18.7- 24.8] |
| Richest | 1513 | 10740 | 14.1 | 25.7 [22.0- 29.5] |
| **Availability of electricity** |  |  |  |  |
| No | 9104 | 10740 | 84.8 | 73.3 [69.5- 77.2] |
| Yes | 1636 | 10740 | 15.2 | 26.7 [22.8- 30.5] |
| **Household's size** |  |  |  |  |
| Median (Q1-Q3) | 6(5-8) |  |  |  |
| 1-3 | 1287 | 10740 | 12.0 | 11.6 [10.4- 12.8] |
| 4-5 | 2757 | 10740 | 25.7 | 27.2 [25.1- 29.3] |
| 6-7 | 3047 | 10740 | 28.4 | 28.5 [26.7- 30.2] |
| ≥ 8 | 3649 | 10740 | 34.0 | 32.8 [30.4- 35.2] |
| **Age of the head of household (year)** |  |  |  |  |
| <30 | 1455 | 10730 | 13.6 | 12.5 [11.4- 13.6] |
| ≥50 | 3700 | 10730 | 34.5 | 37.6 [35.2- 40.0] |
| 30–39 | 2533 | 10730 | 23.6 | 22.7 [21.1- 24.2] |
| 40–49 | 3042 | 10730 | 28.4 | 27.2 [25.6- 28.9] |
| **Sex of the head of household** |  |  |  |  |
| Female | 3325 | 10740 | 31.0 | 33.5 [31.4- 35.6] |
| Male | 7415 | 10740 | 69.0 | 66.5 [64.4- 68.6] |
| **Women age group** |  |  |  |  |
| Median (Q1-Q3) | 27(19-37) |  |  |  |
| 15-19 | 2842 | 10740 | 26.5 | 25.8 [24.5- 27.1] |
| 20-24 | 1931 | 10740 | 18.0 | 18.5 [17.3- 19.7] |
| 25-29 | 1315 | 10740 | 12.2 | 13.2 [12.3- 14.2] |
| 30-34 | 1205 | 10740 | 11.2 | 11.6 [10.7- 12.6] |
| 35-39 | 1286 | 10740 | 12.0 | 12.2 [11.2- 13.1] |
| 40-44 | 1133 | 10740 | 10.5 | 9.8 [9.0- 10.6] |
| 45-49 | 1028 | 10740 | 9.6 | 8.8 [8.0- 9.6] |
| **Education level** |  |  |  |  |
| None | 1872 | 10740 | 17.4 | 15.5 [13.7- 17.3] |
| Primary | 3003 | 10740 | 28.0 | 21.9 [20.2- 23.5] |
| Secondary | 5431 | 10740 | 50.6 | 56.0 [53.8- 58.2] |
| Tertiary | 434 | 10740 | 4.0 | 6.7 [5.2- 8.1] |
| **Marital status** |  |  |  |  |
| Never married | 3736 | 10740 | 34.8 | 39.9 [37.6- 42.2] |
| Married | 3835 | 10740 | 35 | 31.8 [30.0- 33.6] |
| Union | 2021 | 10740 | 18.8 | 18.3 [16.7- 19.9] |
| Widowed | 285 | 10740 | 2.7 | 2.3 [1.9- 2.8] |
| Divorced | 162 | 10740 | 1.5 | 1.5 [1.1- 1.8] |
| Separated | 701 | 10740 | 6.5 | 6.2 [5.4- 6.9] |
| **Number of living children** |  |  |  |  |
| 0 | 3686 | 10740 | 34.3 | 37.9 [36.0- 39.8] |
| 1–2 | 2333 | 10740 | 21.7 | 21.2 [20.0- 22.4] |
| 3–4 | 2088 | 10740 | 19.4 | 19.6 [18.3- 20.8] |
| ≥ 5 | 2633 | 10740 | 24.5 | 21.3 [19.9- 22.7] |
| **Currently working** |  |  |  |  |
| No | 4758 | 10740 | 44.3 | 47.3 [45.1- 49.4] |
| Yes | 5982 | 10740 | 55.7 | 52.7 [50.6- 54.9] |
| **Occupation** |  |  |  |  |
| Not working | 4041 | 10620 | 38.1 | 40.7 [38.5- 42.8] |
| Professional/technical/managerial | 212 | 10620 | 2.0 | 2.2 [1.7- 2.6] |
| Clerical | 76 | 10620 | 0.7 | 1.0 [0.5- 1.4] |
| Agriculture self employed | 3829 | 10620 | 36.1 | 29.3 [26.6- 32.0] |
| Household and domestic | 47 | 10620 | 0.4 | 0.6 [0.4- 0.9] |
| Services | 2111 | 10620 | 19.9 | 22.8 [20.6- 24.9] |
| Skilled manual | 155 | 10620 | 1.5 | 2.2 [1.6- 2.8] |
| Unskilled manual | 149 | 10620 | 1.4 | 1.3 [0.9- 1.6] |
| **Frequency of using internet last month** |  |  |  |  |
| Not at all | 9690 | 10740 | 90.2 | 83.0 [80.7- 85.3] |
| Less than once a week | 125 | 10740 | 1.2 | 1.9 [1.4- 2.5] |
| At least once a week | 264 | 10740 | 2.5 | 4.2 [3.2- 5.1] |
| Almost every day | 661 | 10740 | 6.2 | 10.9 [9.1- 12.7] |
| **Frequency of listening to radio** |  |  |  |  |
| Not at all | 8389 | 10740 | 78.1 | 67.8 [64.8- 70.8] |
| Less than once a week | 944 | 10740 | 8.8 | 10.8 [9.3- 12.2] |
| At least once a week | 1407 | 10740 | 13.1 | 21.4 [19.0- 23.8] |
| **Frequency of watching television** |  |  |  |  |
| Not at all | 7578 | 10740 | 70.6 | 66.9 [64.4- 69.4] |
| Less than once a week | 1594 | 10740 | 14.8 | 17.1 [15.3- 18.8] |
| At least once a week | 1568 | 10740 | 14.6 | 16.0 [14.3- 17.8] |
| **Covered by health insurance** |  |  |  |  |
| No | 10429 | 10740 | 97.1 | 95.6 [94.5- 96.6] |
| Yes | 311 | 10740 | 2.9 | 4.4 [3.4- 5.5] |
| **Currently use contraceptive method** |  |  |  |  |
| No | 9061 | 10740 | 84.4 | 82.9 [81.5- 84.4] |
| Yes | 1679 | 10740 | 15.6 | 17.1 [15.6- 18.5] |
| **Visited health facility during the last 12 months** |  |  |  |  |
| No | 7891 | 10740 | 73.5 | 73.2 [71.2- 75.1] |
| Yes | 2849 | 10740 | 26.5 | 26.8 [24.9- 28.8] |
| **Minimum Dietary diversity Score (MDDS)** |  |  |  |  |
| < 5 groups | 9049 | 10740 | 84.3 | 83.4 [81.7- 85.0] |
| ≥ 5 groups | 1691 | 10740 | 15.7 | 16.6 [15.0- 18.3] |
| **Unhealthy food** |  |  |  |  |
| No | 7021 | 10740 | 65.4 | 61.5 [59.0- 64.0] |
| Yes | 3719 | 10740 | 34.6 | 38.5 [36.0- 41.0] |

**Table 2:**
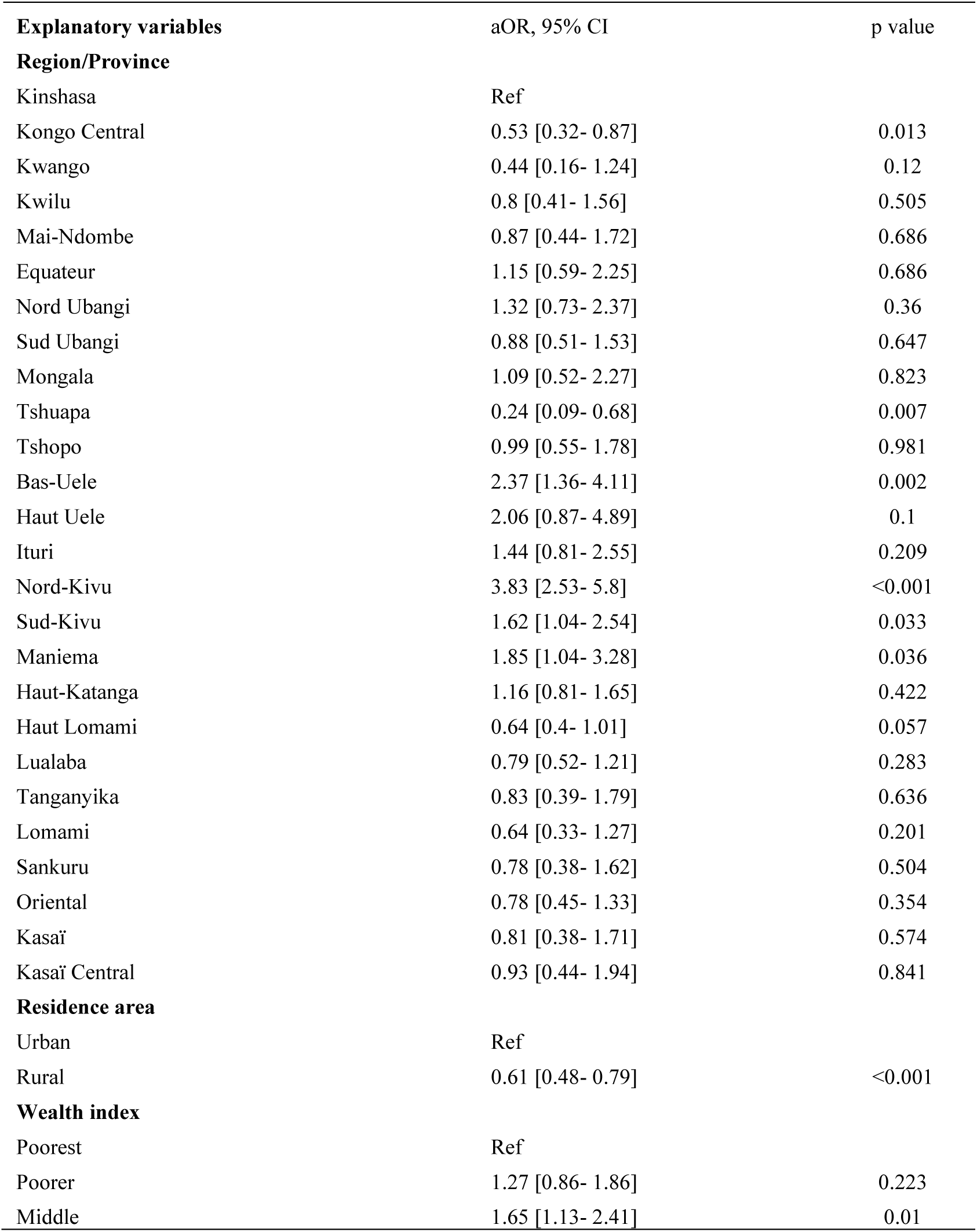
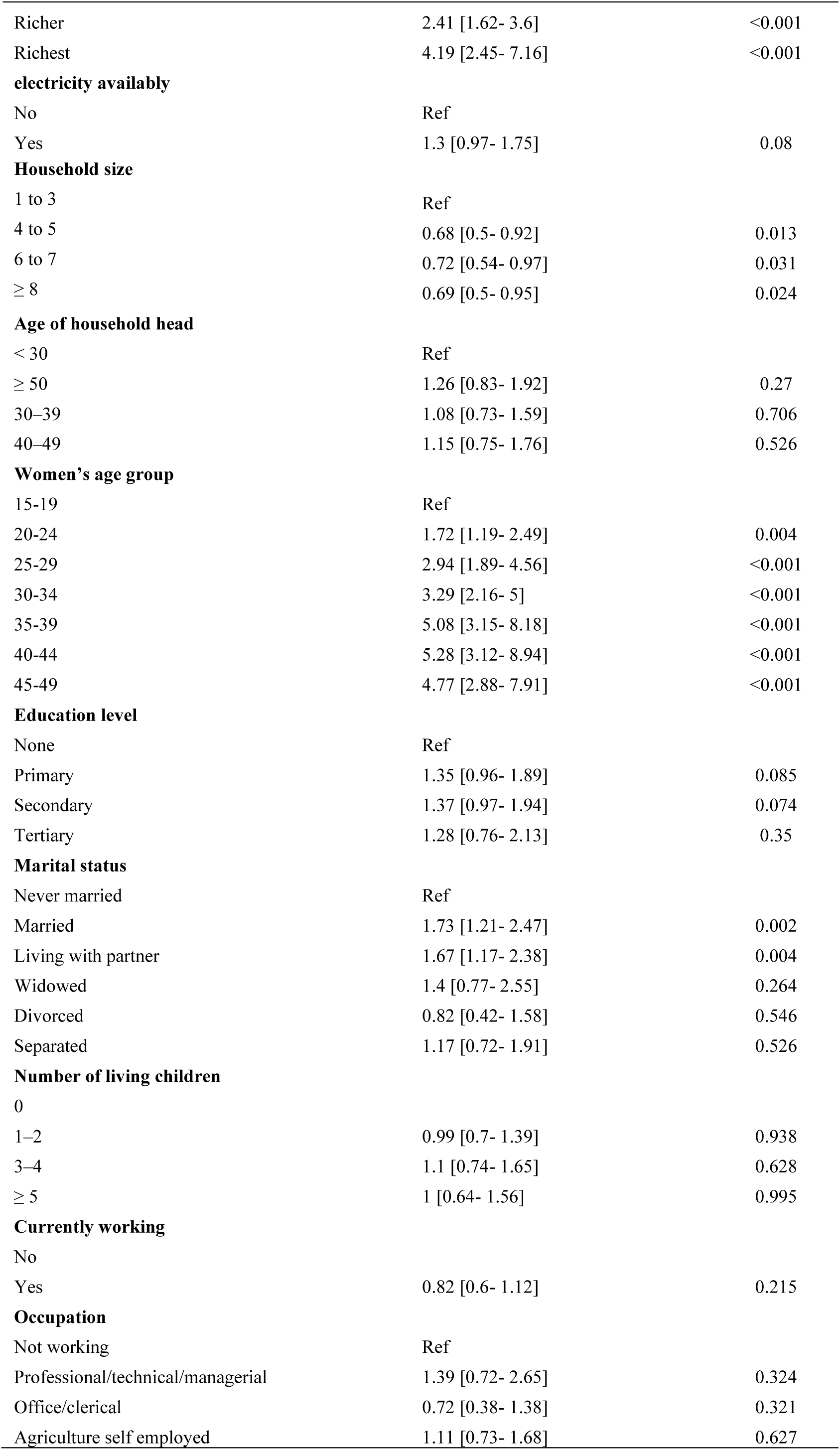
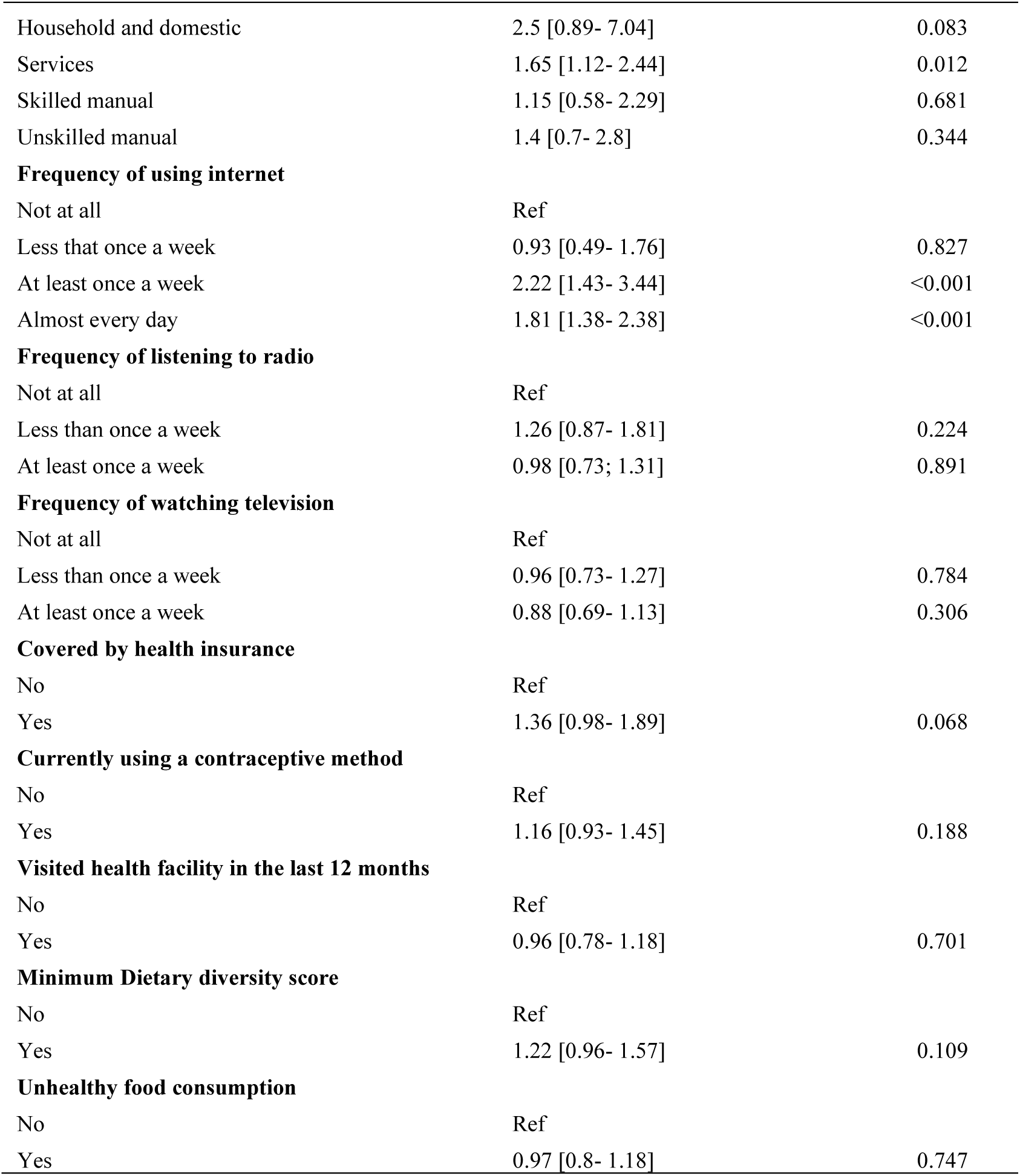
Best fit Multivariate model (IV) for factors associated with overweight/obesity; Supplementary models (I-III) in Annex.

| Explanatory variables | aOR, 95% CI | p value |
| --- | --- | --- |
| **Region/Province** |  |  |
| Kinshasa | Ref |  |
| Kongo Central | 0.53 [0.32- 0.87] | 0.013 |
| Kwango | 0.44 [0.16- 1.24] | 0.12 |
| Kwilu | 0.8 [0.41- 1.56] | 0.505 |
| Mai-Ndombe | 0.87 [0.44- 1.72] | 0.686 |
| Equateur | 1.15 [0.59- 2.25] | 0.686 |
| Nord Ubangi | 1.32 [0.73- 2.37] | 0.36 |
| Sud Ubangi | 0.88 [0.51- 1.53] | 0.647 |
| Mongala | 1.09 [0.52- 2.27] | 0.823 |
| Tshuapa | 0.24 [0.09- 0.68] | 0.007 |
| Tshopo | 0.99 [0.55- 1.78] | 0.981 |
| Bas-Uele | 2.37 [1.36- 4.11] | 0.002 |
| Haut Uele | 2.06 [0.87- 4.89] | 0.1 |
| Ituri | 1.44 [0.81- 2.55] | 0.209 |
| Nord-Kivu | 3.83 [2.53- 5.8] | <0.001 |
| Sud-Kivu | 1.62 [1.04- 2.54] | 0.033 |
| Maniema | 1.85 [1.04- 3.28] | 0.036 |
| Haut-Katanga | 1.16 [0.81- 1.65] | 0.422 |
| Haut Lomami | 0.64 [0.4- 1.01] | 0.057 |
| Lualaba | 0.79 [0.52- 1.21] | 0.283 |
| Tanganyika | 0.83 [0.39- 1.79] | 0.636 |
| Lomami | 0.64 [0.33- 1.27] | 0.201 |
| Sankuru | 0.78 [0.38- 1.62] | 0.504 |
| Oriental | 0.78 [0.45- 1.33] | 0.354 |
| Kasaï | 0.81 [0.38- 1.71] | 0.574 |
| Kasaï Central | 0.93 [0.44- 1.94] | 0.841 |
| **Residence area** |  |  |
| Urban | Ref |  |
| Rural | 0.61 [0.48- 0.79] | <0.001 |
| **Wealth index** |  |  |
| Poorest | Ref |  |
| Poorer | 1.27 [0.86- 1.86] | 0.223 |
| Middle | 1.65 [1.13- 2.41] | 0.01 |
| Richer | 2.41 [1.62- 3.6] | <0.001 |
| Richest | 4.19 [2.45- 7.16] | <0.001 |
| **electricity availably** |  |  |
| No | Ref |  |
| Yes | 1.3 [0.97- 1.75] | 0.08 |
| **Household size** |  |  |
| 1 to 3 | Ref |  |
| 4 to 5 | 0.68 [0.5- 0.92] | 0.013 |
| 6 to 7 | 0.72 [0.54- 0.97] | 0.031 |
| ≥ 8 | 0.69 [0.5- 0.95] | 0.024 |
| **Age of household head** |  |  |
| < 30 | Ref |  |
| ≥ 50 | 1.26 [0.83- 1.92] | 0.27 |
| 30–39 | 1.08 [0.73- 1.59] | 0.706 |
| 40–49 | 1.15 [0.75- 1.76] | 0.526 |
| **Women's age group** |  |  |
| 15-19 | Ref |  |
| 20-24 | 1.72 [1.19- 2.49] | 0.004 |
| 25-29 | 2.94 [1.89- 4.56] | <0.001 |
| 30-34 | 3.29 [2.16- 5] | <0.001 |
| 35-39 | 5.08 [3.15- 8.18] | <0.001 |
| 40-44 | 5.28 [3.12- 8.94] | <0.001 |
| 45-49 | 4.77 [2.88- 7.91] | <0.001 |
| **Education level** |  |  |
| None | Ref |  |
| Primary | 1.35 [0.96- 1.89] | 0.085 |
| Secondary | 1.37 [0.97- 1.94] | 0.074 |
| Tertiary | 1.28 [0.76- 2.13] | 0.35 |
| **Marital status** |  |  |
| Never married | Ref |  |
| Married | 1.73 [1.21- 2.47] | 0.002 |
| Living with partner | 1.67 [1.17- 2.38] | 0.004 |
| Widowed | 1.4 [0.77- 2.55] | 0.264 |
| Divorced | 0.82 [0.42- 1.58] | 0.546 |
| Separated | 1.17 [0.72- 1.91] | 0.526 |
| **Number of living children** |  |  |
| 0 |  |  |
| 1–2 | 0.99 [0.7- 1.39] | 0.938 |
| 3–4 | 1.1 [0.74- 1.65] | 0.628 |
| ≥ 5 | 1 [0.64- 1.56] | 0.995 |
| **Currently working** |  |  |
| No |  |  |
| Yes | 0.82 [0.6- 1.12] | 0.215 |
| **Occupation** |  |  |
| Not working | Ref |  |
| Professional/technical/managerial | 1.39 [0.72- 2.65] | 0.324 |
| Office/clerical | 0.72 [0.38- 1.38] | 0.321 |
| Agriculture self employed | 1.11 [0.73- 1.68] | 0.627 |
| Household and domestic | 2.5 [0.89- 7.04] | 0.083 |
| Services | 1.65 [1.12- 2.44] | 0.012 |
| Skilled manual | 1.15 [0.58- 2.29] | 0.681 |
| Unskilled manual | 1.4 [0.7- 2.8] | 0.344 |
| **Frequency of using internet** |  |  |
| Not at all | Ref |  |
| Less than once a week | 0.93 [0.49- 1.76] | 0.827 |
| At least once a week | 2.22 [1.43- 3.44] | <0.001 |
| Almost every day | 1.81 [1.38- 2.38] | <0.001 |
| **Frequency of listening to radio** |  |  |
| Not at all | Ref |  |
| Less than once a week | 1.26 [0.87- 1.81] | 0.224 |
| At least once a week | 0.98 [0.73; 1.31] | 0.891 |
| **Frequency of watching television** |  |  |
| Not at all | Ref |  |
| Less than once a week | 0.96 [0.73- 1.27] | 0.784 |
| At least once a week | 0.88 [0.69- 1.13] | 0.306 |
| **Covered by health insurance** |  |  |
| No | Ref |  |
| Yes | 1.36 [0.98- 1.89] | 0.068 |
| **Currently using a contraceptive method** |  |  |
| No | Ref |  |
| Yes | 1.16 [0.93- 1.45] | 0.188 |
| **Visited health facility in the last 12 months** |  |  |
| No | Ref |  |
| Yes | 0.96 [0.78- 1.18] | 0.701 |
| **Minimum Dietary diversity score** |  |  |
| No | Ref |  |
| Yes | 1.22 [0.96- 1.57] | 0.109 |
| **Unhealthy food consumption** |  |  |
| No | Ref |  |
| Yes | 0.97 [0.8- 1.18] | 0.747 |

### Prevalence of overweight and obesity

Among the 10740 women included in the analysis, the weighted prevalence of overweight wa s 13.3(95%CI: 12-14.7), while obesity affected 6.2(95%CI: 5.2-7.2). When combining both conditions, the pooling prevalence of overweight or obesity reached 19.5 (95% CI: 17.6-21.4).

### Sensitivity analysis 15-49 vs 20-49 years

Restricting the sample to women aged 20-49 years increased the national prevalence of overweight/obesity to 23.7% (95% CI: 21.3-26.1). Prevalence among adolescents aged 15-49 years was substantially lower (7.5%, 95% CI: 5.7-9.3), resulting in absolute difference of −4.2 percentage points between the 15-49 and 20-49 estimates, consistent with an age composition dilution effect. Urban rural disparities remained pronounced under in both specifications: 31.6% vs. 10.1% (15-49 years) and 40 % vs 11.6% (20-49 years).

### Bivariate analysis

In Bivariate analysis, several variables met the inclusion criterion (p<0.2) for entry into the multivariable model. These include geographic region, residence area, household wealth index, electricity access, household size, and age of the household head. Women’s age group, education level, marital status, number of living children, employment and occupation, as well media exposure, health insurance coverage, contraceptive use m recent health facility attendance, dietary diversity, and unhealthy food consumption were also retained (Annex 1).

### Multivariate analysis

Successive multivariable models were developed, beginning with core sociodemographic vari ables and progressively adding households, economic, and behavioural factors. Model IV, rep resenting the fully adjusted specification and the best performing model, is reporting in this section. It’s the most comprehensive model (Table 3).

### Region

Adjusted odds ratios varied substantially across provinces when compared with Kinshasa region. Lower estimates were observed in Kongo Central (aOR=0.53; 95%CI:0.32-0.87), Tshuapa (aOR=0.24;0.09-0.68), and Haut-Lomami (aOR=0.64;0.4-1.01).

Higher adjusted odds ratios were observed in Bas-Uele (aOR=2.37;95%CI: 1.36-4.11), Nord-Kivu (aOR=3.83;95%CI:2.53-5.8), Sud-Kivu (aOR =1.62;95%CI: 1.04-2.54), and Maniema (aOR=1.85;95%CI: 1.04-3.28).

### Residence and household characteristics

Women living in rural areas had lower adjusted odds than those in urban areas (aOR =0.61;95%CI:0.48-0.79). Household wealth index status showed a graded pattern, with increasing adjusted odds across the middle (aOR=1.65; 1.13-2.41), richer (aOR=2.41;1.62-3.60), and richest wealth categories (aOR=4.19;2.45-7.16), compared with the poorest households. Electricity availability did not reach statistical significance (aOR =1.30;0.97-1.75). Larger households showed lower adjusted odds relative to those with 1-3 members; 4-5 members (aOR =0.68;0.50-0.92), 6-7 members (aOR =0.72;0.54-0.97), and ≥ 8 members (aOR =0.69;0.5-0.95).

### Wealth index

A clear sociodemographic gradient was observed across the household wealth index. Compared with women from the poorest households, adjusted odds ratios increased possessively across the wealth distribution. Estimates were slightly higher in the poorer category (aOR=1.27; 95% CI: 0.86-1.86), followed by a more marked increase in the middle group (aOR=1.65; 95% CI: 1.13-2.41). Higher values were observed in the richer households (aOR=12.41; 1.62-3.60), with the highest estimate recorded among women in the richest households (aOR=14.19; 2.45-7.16).

### Sociodemographic factors

A strong age-related pattern was observed. Relative to adolescents aged 15-19 years, adjusted odds increased steadily from early adulthood 9 20-24 years; aOR=1.72;95%CI: 1.19-2.49) through ages 25-29 (aOR=2.94; 95%CI: 1.89-4.56) and 30-34 (3.29;2,16-5). The highest estimates were observed among women aged 35-39 (aOR=5.08; 95% CI: 3.15-8,18) and 40-44 years (aOR=5.28; 95%CI: 3.12-8.94). Among the oldest group (45-49 years), adjusted odds remained elevated (aOR=4.77;95%CI: 2.88-7.91) compared with the youngest category.

Educational attainment did not show statistically significant differences, with overlapping confidence across primary (aOR=1.35; 95%CI: 0.95-1.89), secondary (aOR=1,37;95%CI =0.97.194), and tertiary levels (aOR=1.28; 95 CI: 0.76-2.13).

Marital status showed variation across categories. Compared with never marries women, higher adjusted odd was observed among married women (aOR =1.73;95%CI: 1.21-2.47) and those living with a partner (aOR=1.67;95%CI: 1,17-2.38). Estimates among widowed (aOR=1.4;95%CI: 0.77-2.55), divorces (aOR=0.82; 95%CI: 0.42-1.58), and separated women (aOR=1.17;95%CI: 0.72-1.91) did not reach statistical significance.

### Occupation

Across occupational categories, estimates varied but were mostly non-significant. Relative to women not working, adjusted odds ratio was (aOR=1.39; 95%CI: 0.72-2.65) for professional or technical occupations, (aOR =0.72; 95%CI: 0.38-1.38) for office/clerical roles, (aOR =1.11;95%CI: 0.73-1.68) for self-employed agricultural workers, and (aOR=1.15;95%CI: 058-2.29) for skilled manual workers. The only category showing higher adjusted odds was service occupations (aOR=1.65; 95%CI:1.12-2.44). Unskilled manual work did not show statistically significant differences (aOR=1.40;95%CI: 0.7-2.80).

### Media exposure

Patterns differ across media types. Internet use was associated with higher adjusted odds among women reporting use at least once a week aOR=2. 23;95%CI: 1.43-3.44) and almost daily (aOR=1,91;95%CI: 1.38-2.38). compared with non-users, no statistical differences were observed for radio listening or television viewing across their respective categories.

### Health related and dietary factors

Health insurance coverage, contraceptive use and recent health facility visit did not show significant differences in the adjusted model. Similarly, neither minimum dietary diversity nor unhealthy food consumption demonstrated statistically significant associations.

## Discussion

Using data from the 2023 Demographic and Health survey (DHS) in the DRC, this study estimates the prevalence of overweight/obesity among women of reproductive age and explore their distribution across contextual, household, sociodemographic and behavioural factors. The nationally weighted prevalence was 19.53%, which remains lower than pooled estimates reported for Sub Saharan Africa but is substantially higher in urban areas (31.6%), consistent with patterns reported in comparable African settings(1,2,20). Notably, this estimate represents a marked increase compared with 2013-14 DHS, where the prevalence of overweight and obesity among women was estimated at 15% indicating a rapid rise over the past decade (21). To preserve comparability with national DHS reporting and prior DHS based literature, the primary analysis used the standard 15-49 years range. In our data, adolescents aged 15-19 years age range. In our data, adolescents aged 15-199 years represented roughly one quarter of the analytic sample and had markedly lower overweight/obesity prevalence (7.5%), which partly explains the lower overall estimates for 15-49 years compared with 20-49 restricted sample (23.7%). Importantly, sensitivity analyses indicated that key structural gradients, particularly the strong urban-rural disparity remained substantial after excluding adolescents, suggesting that inferences on contextual and socioeconomic determinants are unlikely to be driven by adolescent inclusion or age-related misclassification. Moreover, Comparisons of aOR from the fully adjusted model across the 15-49 and 20-40 (Anex 11) indicated that core association as residence or wealth index were generally stable, strengthening the robustness of the of findings. Prior to the 2023 DHS, local studies in the DRC had already documented heterogeneity in overweight and obesity across age groups and settings. In south Kivu, a community based study among adults aged 18 years and older reports a hight combined prevalence of 33.6%, including 7.1% obesity), whereas among university students in Kinshasa, excess weight was more common among women than men (13.4 versus 9%) (23,24). Together, these findings suggested an emerging burden of excess weight well before it became apparent in nationally representative data.

Our results reveal substantial socioeconomic and demographic inequalities in overweight and obesity among women in the DRC, reflecting broader patterns of the nutrition transition observed across Sub-Saharan Africa and other low and middle income countries (6,10,16,20). Marked geographical disparities were observed, with Kinshasa remaining among the provinces with the highest prevalence, estimated at 38% (32.8-43.2). Women residing in several eastern provinces, Nord Kivu, Bas Uele and Maniema had substantially higher odds of overweight/obesity, whereas lower odds were observed in provinces such as Tshuapa and Kongo Central. Theses contrasts may reflect heterogeneous stages of nutritional within the country.

Similar subnational disparities have been reported in countries such as Nigeria, Ethiopia and Tanzania where variation in urbanization, food systems and economic development contribute to uneven distributions of overweight/obesity. In conflict affected settings, population displacement, disrupted food market and increased reliance on inexpensive energy dense foods may accelerate transitions towards overweight and obesity among women(15,17,18,30).

The observed urban-rural and wealth gradients are consistent with extensive evidence from Sub Saharan Africa showing that overweight and obesity disproportionally affect urban and wealthier populations(15,16). For instance, DHS based analysis in Ghana, Kenya and South Africa have reported similar monotonic increased risk across wealth quintiles, reflecting differential exposure to obesogenic environments(16,19).These environments are characterised by greater access to ultra proceed and energy dense foods, reduced physical occupational and transport related physical activity, and a progressive shift toward sedentary lifestyles accompanying socioeconomic advancement (6,19,20).

Beyond Africa, evidence South Asia and Latin America also demonstrates similar socio-economic gradients. However, in several middle-income countries, the burden of overweight and obesity is increasingly shifting toward poorer urban populations, highlighting a more advanced stage of nutrition transition (20). In this context, the persistence of higher prevalence among women living in wealthier household in DRC suggests that the country remains at an earlier stage of this nutritional transition.

Age emerged as one of the strongest correlates of overweight and obesity, with markedly higher odds among women aged 30 year and older. This finding is consistent with multi country evidence, and may reflect cumulative life course exposure to behavioural and metabolic risk factors (11). Biological and sociocultural mechanisms may also contribute this pattern, including pregnancy-related weight retention, parity related metabolic changes, evolving household dietary roles, sociocultural norms that valorise larger body size, and increasing constraints on time available for physical activity as women age. (13,16,20,27,28).

In contrast, education level, employment status, and several behavioural indicators, including dietary diversity and unhealthy food consumption, were not independently associated with overweight/obesity. Similar null or inconsistent associations have been reported in DHS based studies from west and central Africa, highlighting the limitations of cross-sectional dietary proxies and the difficulty of capturing complex food environments and energy balance using standard survey instruments. The positive association with frequent internet use, but not television or radio exposure, suggests emerging lifestyle patterns linked to digital access and sedentary behaviour a phenomenon increasingly observed in urban African contexts.

### Implications for public health, policy and research

These findings highlight clear priorities for action int the Democratic Republic of Congo. From a public health perspective, efforts to prevent overweight and obesity should prioritise urban and wealthier populations, where le burden is currently concentrated. While structural interventions addressing food environments, urban design, transport systems and access to healthy foods are essential for sustainable impact; they must be complemented by context including cultural appropriate health promotion and community-based sensitisation that fit with women’s daily realities. Together, these approaches directly support Sustainable Development Goal (SGD) 3 (ensure healthy lives and promote wellbeing for all) and SGD 11 (make cities inclusive, safe and sustainable).

The pronounced provincial disparities observed underscore the need for sub-national tailored strategies, that combine national policy actions with locally adapted communication delivered through primary health services, community platforms and women focused programmes. Without strong multisectoral food, urban and transport policies, the burden of excess weight is likely to progressively shift toward poorer populations, as seen in more advanced stages of nutrition transition.

To effectively curb the growing burden of overweight and obesity in the DRC, WHO “Best Buys” and UNICEF-WHO double duty must be implemented to reduce current urban inequalities and prevent the future shift(25,31).

From a research perspective, strengthening longitudinal and improving the measurement of diet, physical activity and exposure to health promotion within national surveys would better inform equitable and effective responses.

### Strengths and limitations

This study has several strengths, including the use of nationally representative DHS data, standardized anthropometric measurements, and survey weighted analyses accounting for the complex design, which enable robust subnational comparisons.

However, several limitations should be acknowledged. The cross-sectional design limits causal inference. Dietary diversity and unhealthy food consumption indicators are based on short recall periods and may not adequately capture habitual intake or total energy consumption(32,33). Physical activity was not directly measured, limiting interpretation of behavioural pathways. Finally, although DHS data are highly standardized, residual confounding related to unmeasured of some contextual factors, such as local food environments or insecurity, may persist and should be considered when interpreting the findings.

## List of abbreviations

aOR: Adjusted odds ratio
CI: Confidence interval
DBM: Double burden of malnutrition
DHS: Demographic and Health Survey
DRC: Democratic Republic of the Congo
IR: Individual record
OR: Odd ratio
WHO: World Health Organization

## Author contributions

SB conceptualized the study and formulated the research questions. SB and XX assessed the data, performed the statistical analysis, interpreted the results and wrote the draft of the report. (PM, SJ, BM, DMK) Critically reviewed the manuscript. All the authors approved the final version.

## Acknowledgments

We acknowledge the DHS program for providing access to the 2023 DRCongo DHS data used in this study.

## Consent to publish declaration

Not applicable.

## Data availability statement

The authors used data from the Demographic and Health Surveys, which are publicly available and can be accessed from https://dhsprogram.com.

## Supplementary material

**Annex 1:**
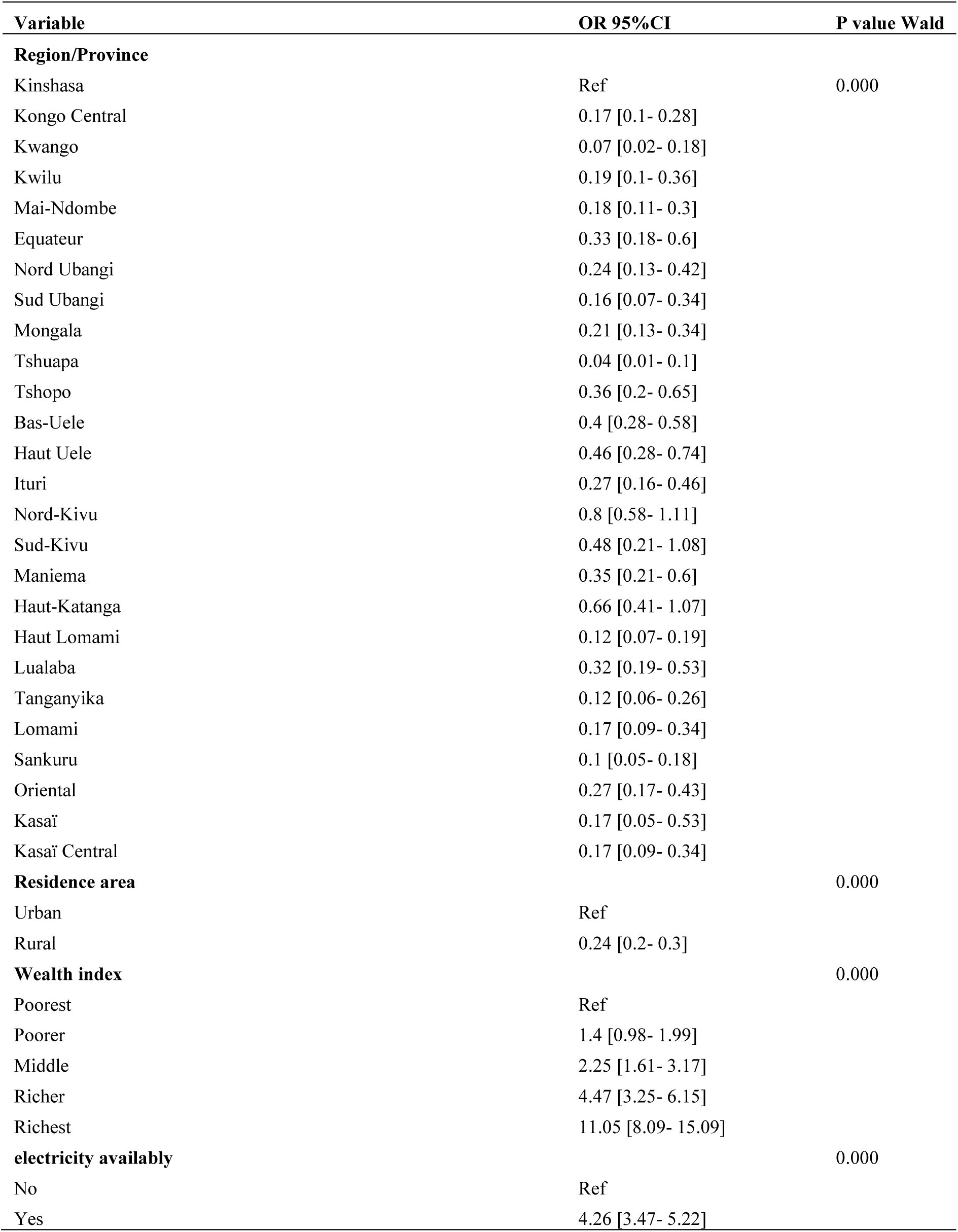
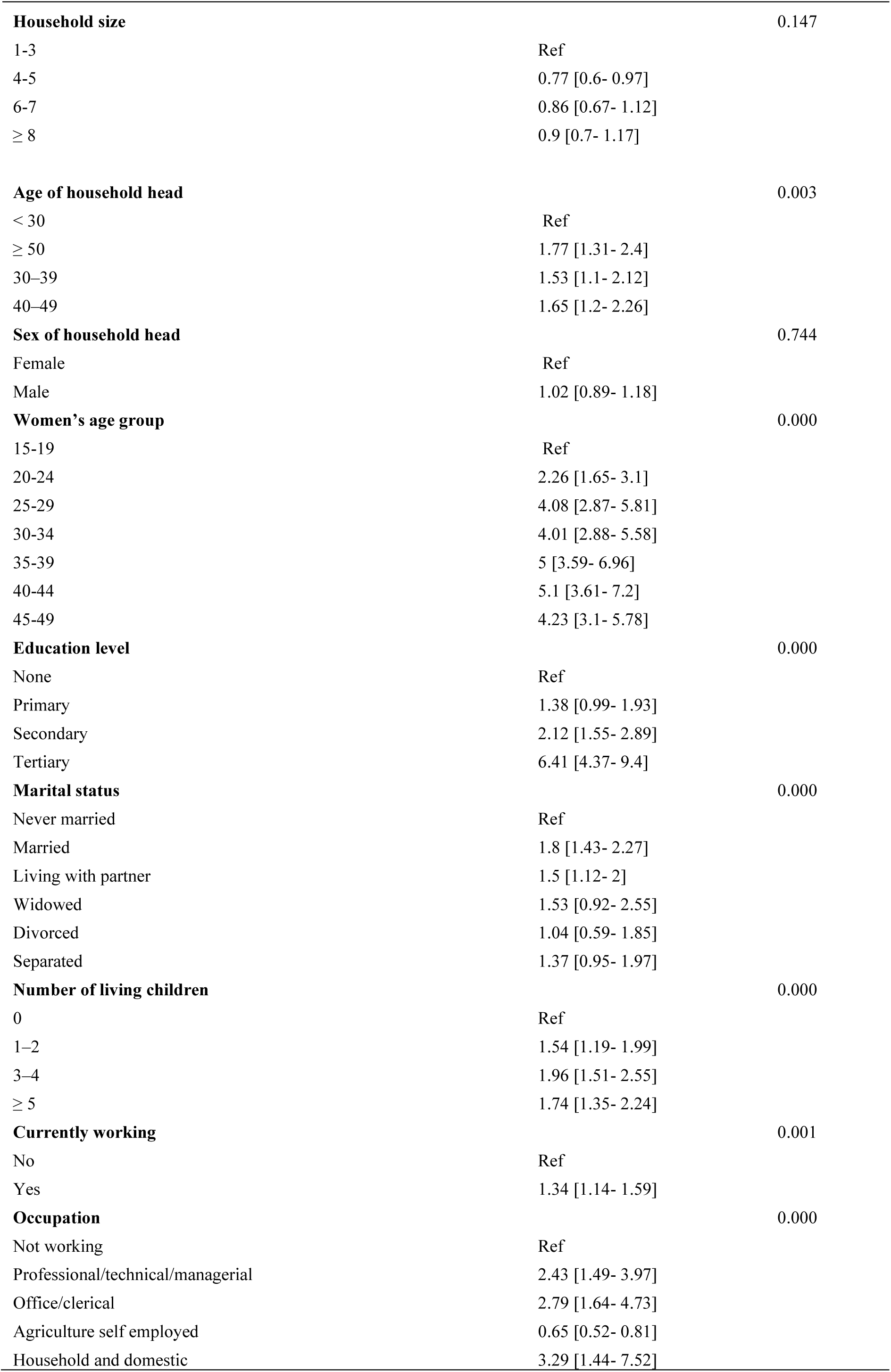
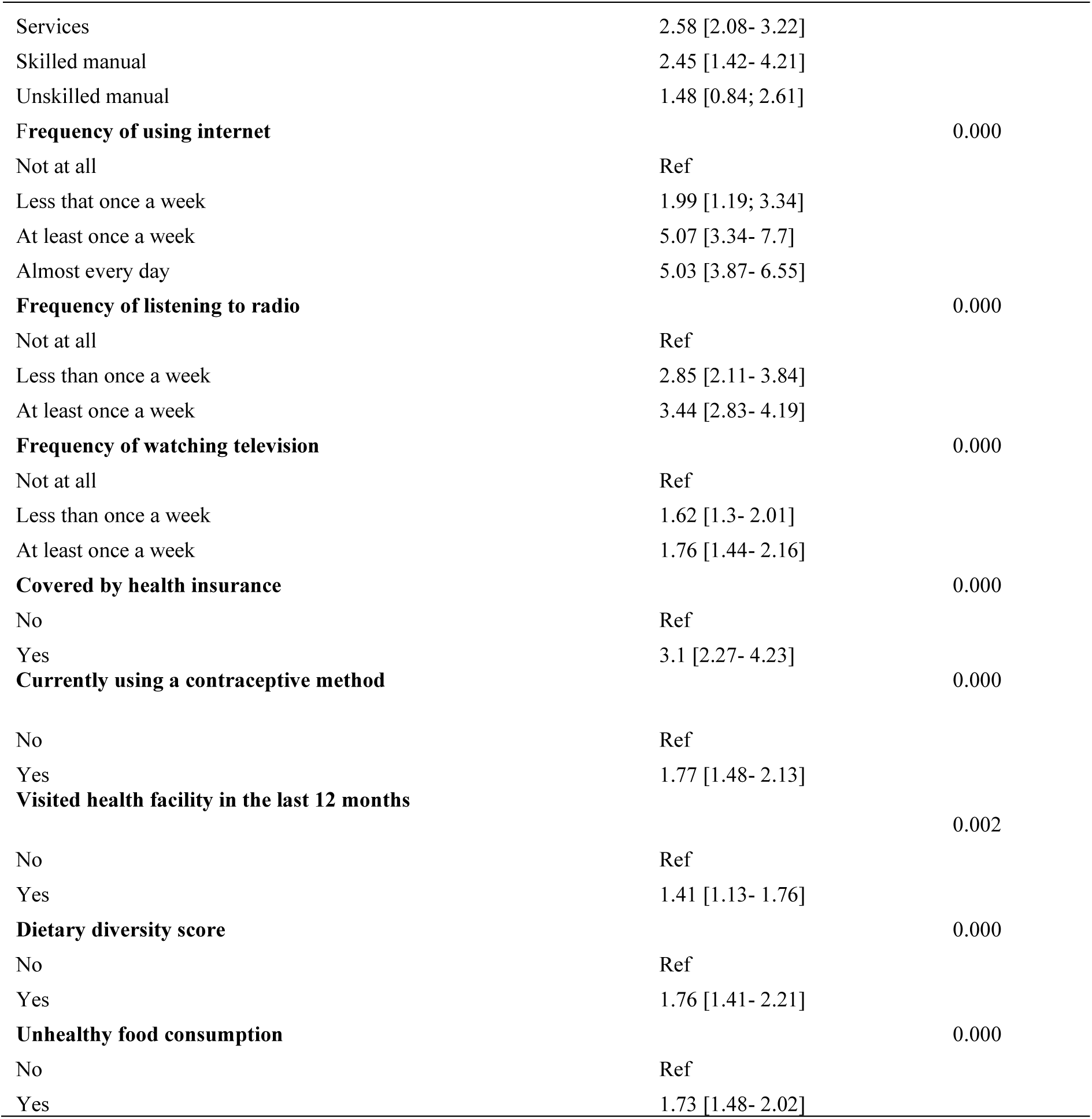
Bivariate analysis of factors associated with overweight/obesity.

| Variable | OR 95%CI | P value Wald |
| --- | --- | --- |
| **Region/Province** |  |  |
| Kinshasa | Ref | 0.000 |
| Kongo Central | 0.17 [0.1- 0.28] |  |
| Kwango | 0.07 [0.02- 0.18] |  |
| Kwilu | 0.19 [0.1- 0.36] |  |
| Mai-Ndombe | 0.18 [0.11- 0.3] |  |
| Equateur | 0.33 [0.18- 0.6] |  |
| Nord Ubangi | 0.24 [0.13- 0.42] |  |
| Sud Ubangi | 0.16 [0.07- 0.34] |  |
| Mongala | 0.21 [0.13- 0.34] |  |
| Tshuapa | 0.04 [0.01- 0.1] |  |
| Tshopo | 0.36 [0.2- 0.65] |  |
| Bas-Uele | 0.4 [0.28- 0.58] |  |
| Haut Uele | 0.46 [0.28- 0.74] |  |
| Ituri | 0.27 [0.16- 0.46] |  |
| Nord-Kivu | 0.8 [0.58- 1.11] |  |
| Sud-Kivu | 0.48 [0.21- 1.08] |  |
| Maniema | 0.35 [0.21- 0.6] |  |
| Haut-Katanga | 0.66 [0.41- 1.07] |  |
| Haut Lomami | 0.12 [0.07- 0.19] |  |
| Lualaba | 0.32 [0.19- 0.53] |  |
| Tanganyika | 0.12 [0.06- 0.26] |  |
| Lomami | 0.17 [0.09- 0.34] |  |
| Sankuru | 0.1 [0.05- 0.18] |  |
| Oriental | 0.27 [0.17- 0.43] |  |
| Kasaï | 0.17 [0.05- 0.53] |  |
| Kasaï Central | 0.17 [0.09- 0.34] |  |
| **Residence area** |  | 0.000 |
| Urban | Ref |  |
| Rural | 0.24 [0.2- 0.3] |  |
| **Wealth index** |  | 0.000 |
| Poorest | Ref |  |
| Poorer | 1.4 [0.98- 1.99] |  |
| Middle | 2.25 [1.61- 3.17] |  |
| Richer | 4.47 [3.25- 6.15] |  |
| Richest | 11.05 [8.09- 15.09] |  |
| **electricity availably** |  | 0.000 |
| No | Ref |  |
| Yes | 4.26 [3.47- 5.22] |  |
| **Household size** |  | 0.147 |
| 1-3 | Ref |  |
| 4-5 | 0.77 [0.6- 0.97] |  |
| 6-7 | 0.86 [0.67- 1.12] |  |
| ≥ 8 | 0.9 [0.7- 1.17] |  |
| **Age of household head** |  | 0.003 |
| < 30 | Ref |  |
| ≥ 50 | 1.77 [1.31- 2.4] |  |
| 30–39 | 1.53 [1.1- 2.12] |  |
| 40–49 | 1.65 [1.2- 2.26] |  |
| **Sex of household head** |  | 0.744 |
| Female | Ref |  |
| Male | 1.02 [0.89- 1.18] |  |
| **Women's age group** |  | 0.000 |
| 15-19 | Ref |  |
| 20-24 | 2.26 [1.65- 3.1] |  |
| 25-29 | 4.08 [2.87- 5.81] |  |
| 30-34 | 4.01 [2.88- 5.58] |  |
| 35-39 | 5 [3.59- 6.96] |  |
| 40-44 | 5.1 [3.61- 7.2] |  |
| 45-49 | 4.23 [3.1- 5.78] |  |
| **Education level** |  | 0.000 |
| None | Ref |  |
| Primary | 1.38 [0.99- 1.93] |  |
| Secondary | 2.12 [1.55- 2.89] |  |
| Tertiary | 6.41 [4.37- 9.4] |  |
| **Marital status** |  | 0.000 |
| Never married | Ref |  |
| Married | 1.8 [1.43- 2.27] |  |
| Living with partner | 1.5 [1.12- 2] |  |
| Widowed | 1.53 [0.92- 2.55] |  |
| Divorced | 1.04 [0.59- 1.85] |  |
| Separated | 1.37 [0.95- 1.97] |  |
| **Number of living children** |  | 0.000 |
| 0 | Ref |  |
| 1–2 | 1.54 [1.19- 1.99] |  |
| 3–4 | 1.96 [1.51- 2.55] |  |
| ≥ 5 | 1.74 [1.35- 2.24] |  |
| **Currently working** |  | 0.001 |
| No | Ref |  |
| Yes | 1.34 [1.14- 1.59] |  |
| **Occupation** |  | 0.000 |
| Not working | Ref |  |
| Professional/technical/managerial | 2.43 [1.49- 3.97] |  |
| Office/clerical | 2.79 [1.64- 4.73] |  |
| Agriculture self employed | 0.65 [0.52- 0.81] |  |
| Household and domestic | 3.29 [1.44- 7.52] |  |
| Services | 2.58 [2.08- 3.22] |  |
| Skilled manual | 2.45 [1.42- 4.21] |  |
| Unskilled manual | 1.48 [0.84; 2.61] |  |
| **Frequency of using internet** |  | 0.000 |
| Not at all | Ref |  |
| Less than once a week | 1.99 [1.19; 3.34] |  |
| At least once a week | 5.07 [3.34- 7.7] |  |
| Almost every day | 5.03 [3.87- 6.55] |  |
| **Frequency of listening to radio** |  | 0.000 |
| Not at all | Ref |  |
| Less than once a week | 2.85 [2.11- 3.84] |  |
| At least once a week | 3.44 [2.83- 4.19] |  |
| **Frequency of watching television** |  | 0.000 |
| Not at all | Ref |  |
| Less than once a week | 1.62 [1.3- 2.01] |  |
| At least once a week | 1.76 [1.44- 2.16] |  |
| **Covered by health insurance** |  | 0.000 |
| No | Ref |  |
| Yes | 3.1 [2.27- 4.23] |  |
| **Currently using a contraceptive method** |  | 0.000 |
| No | Ref |  |
| Yes | 1.77 [1.48- 2.13] |  |
| **Visited health facility in the last 12 months** |  | 0.002 |
| No | Ref |  |
| Yes | 1.41 [1.13- 1.76] |  |
| **Dietary diversity score** |  | 0.000 |
| No | Ref |  |
| Yes | 1.76 [1.41- 2.21] |  |
| **Unhealthy food consumption** |  | 0.000 |
| No | Ref |  |
| Yes | 1.73 [1.48- 2.02] |  |

**Annex 2 :**
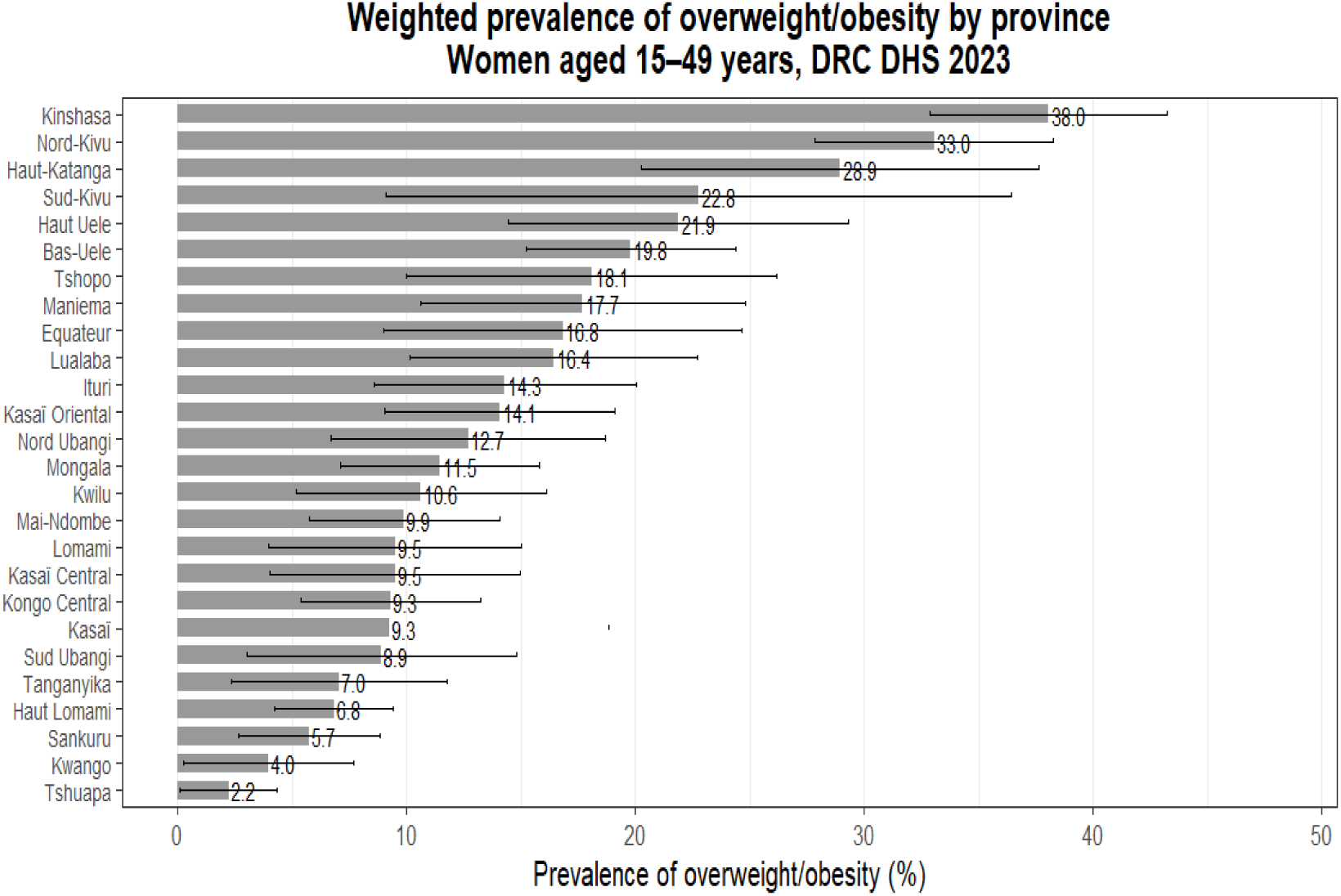
Weighted prevalence of overweight/obesity among women aged 15-49 years by province.

**Annex 3 :**
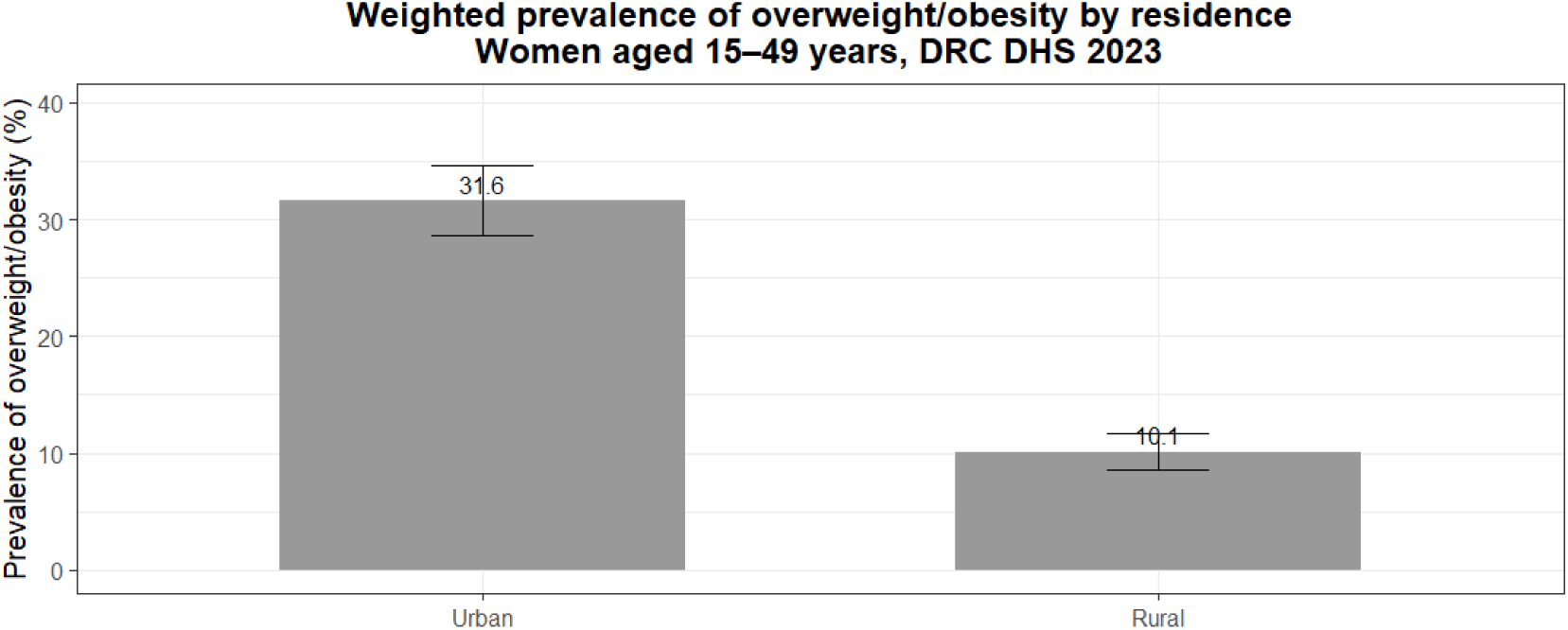
Weighted prevalence (%) of overweight/obesity among women aged 15-49 years by prevalence.

**Annex 4:**
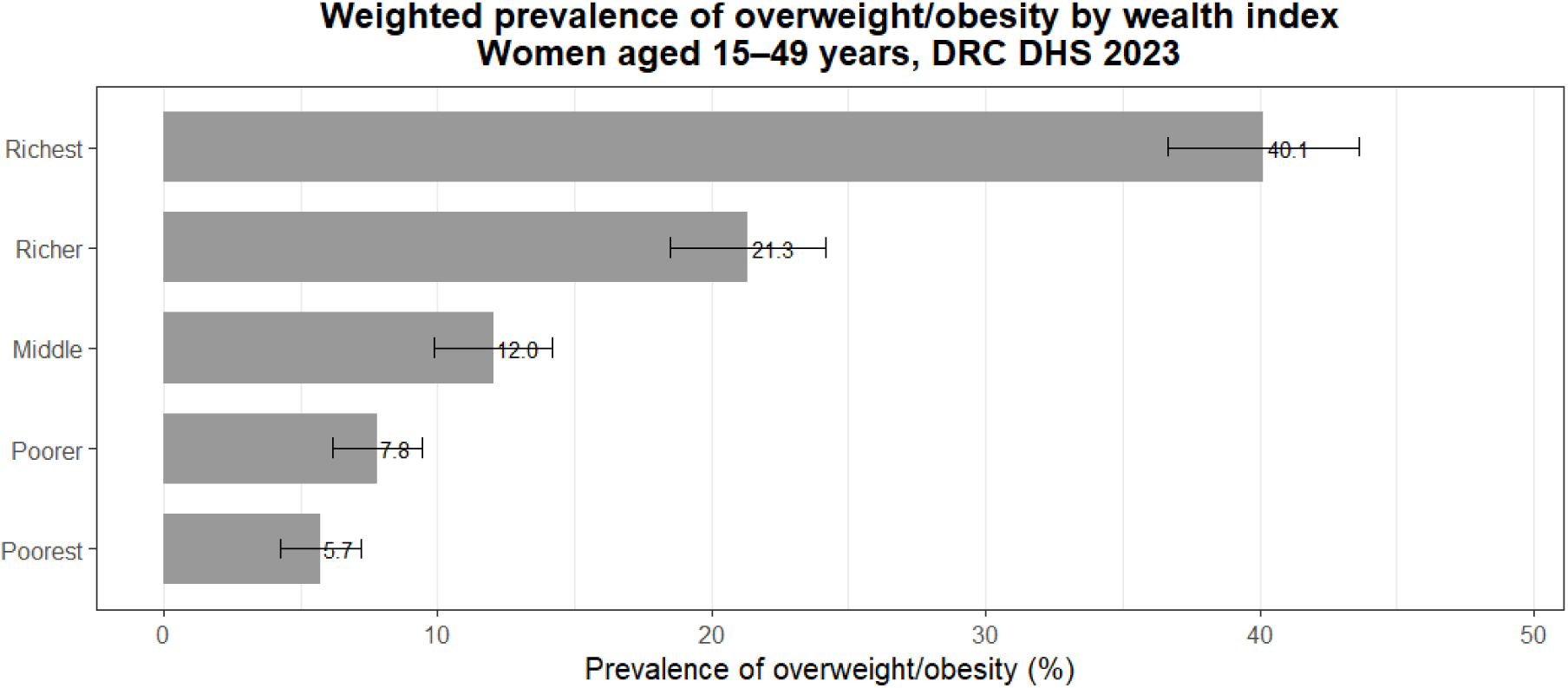
Weighted prevalence of overweight/obesity among women aged 15-49 years by wealth index.

**Annex 5:**
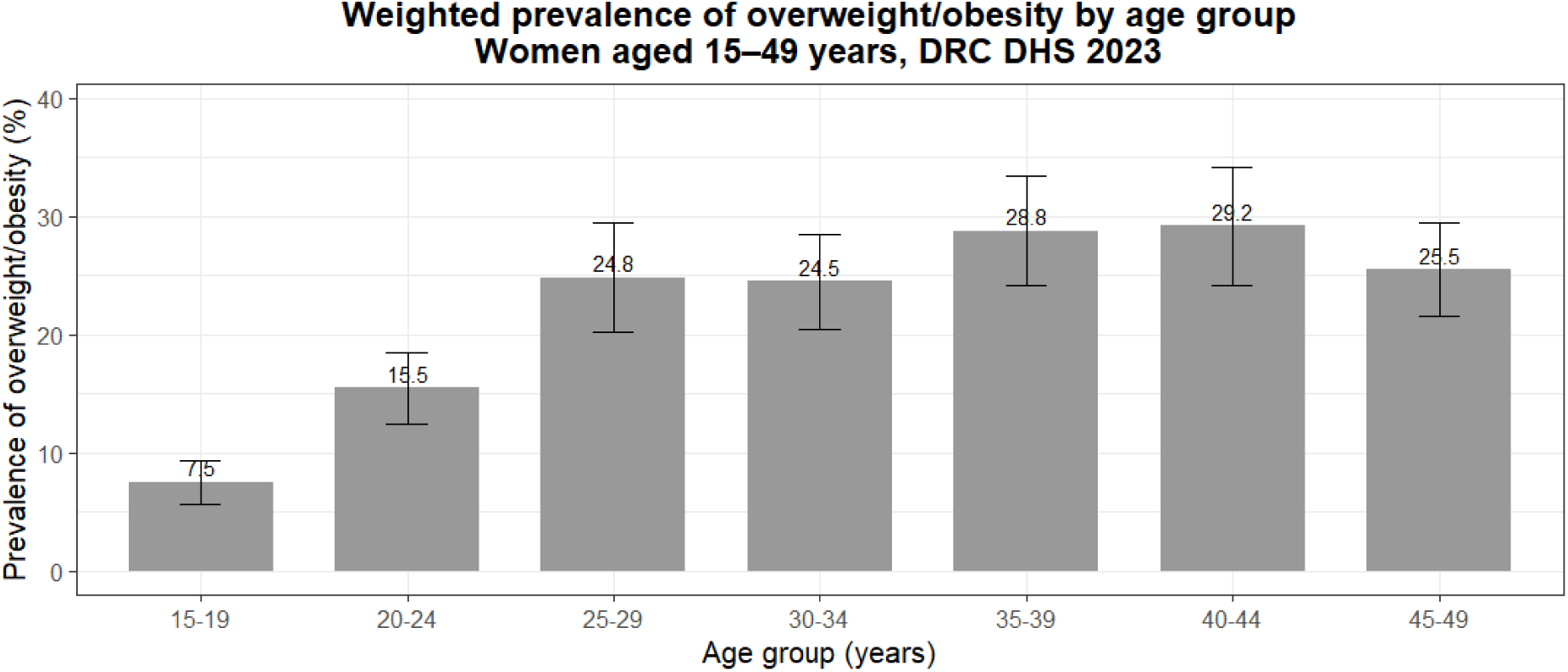
Weighted prevalence of overweight/obesity among women aged 15-49 years by height age group.

**Annex 6 :**
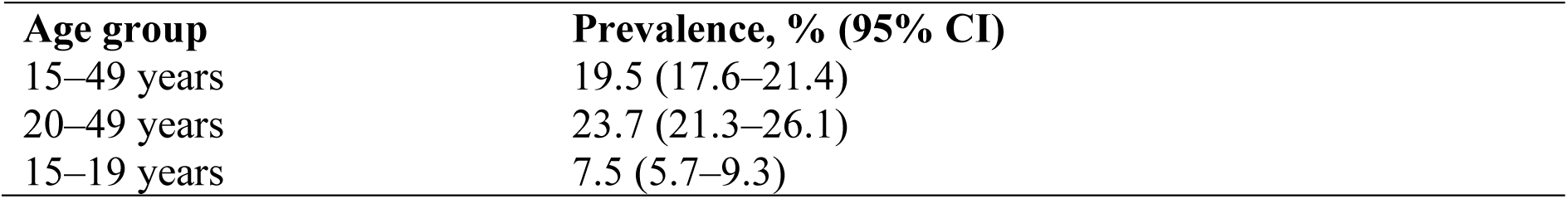
Weighted prevalence of overweight/obesity for sensitivity analysis.

| Age group | Prevalence, % (95% CI) |
| --- | --- |
| 15–49 years | 19.5 (17.6–21.4) |
| 20–49 years | 23.7 (21.3–26.1) |
| 15–19 years | 7.5 (5.7–9.3) |

**Annex 7 :**
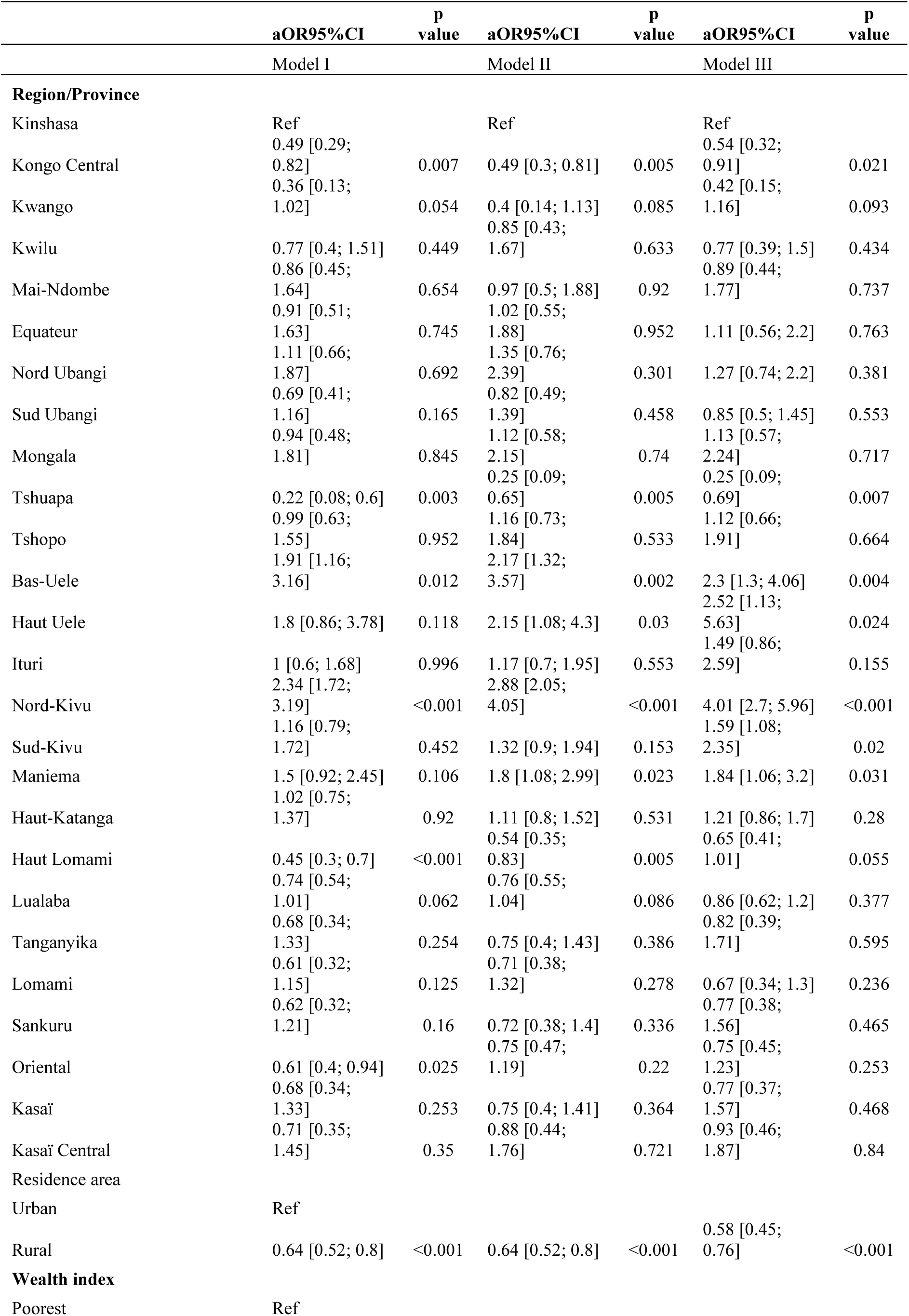
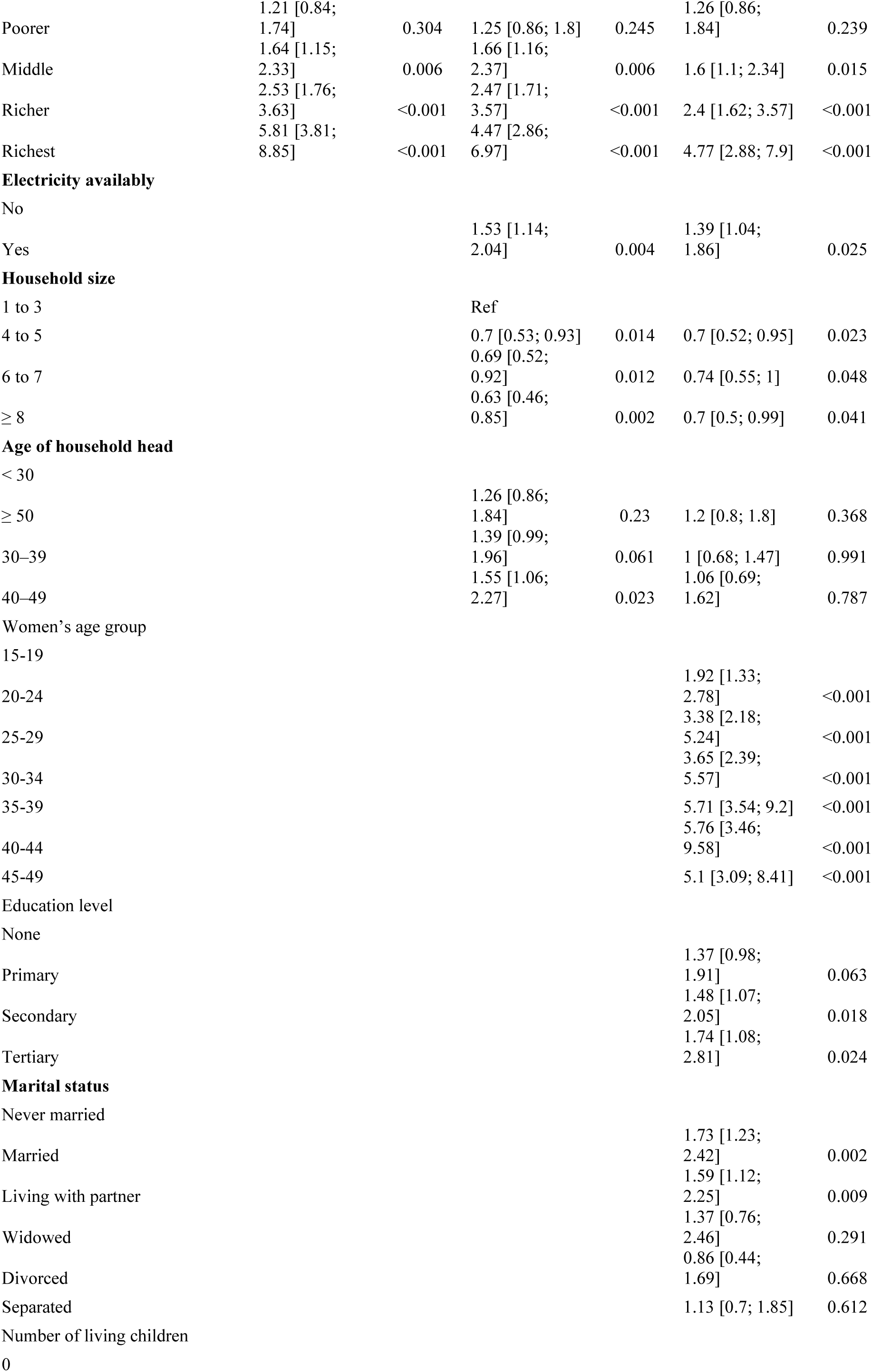
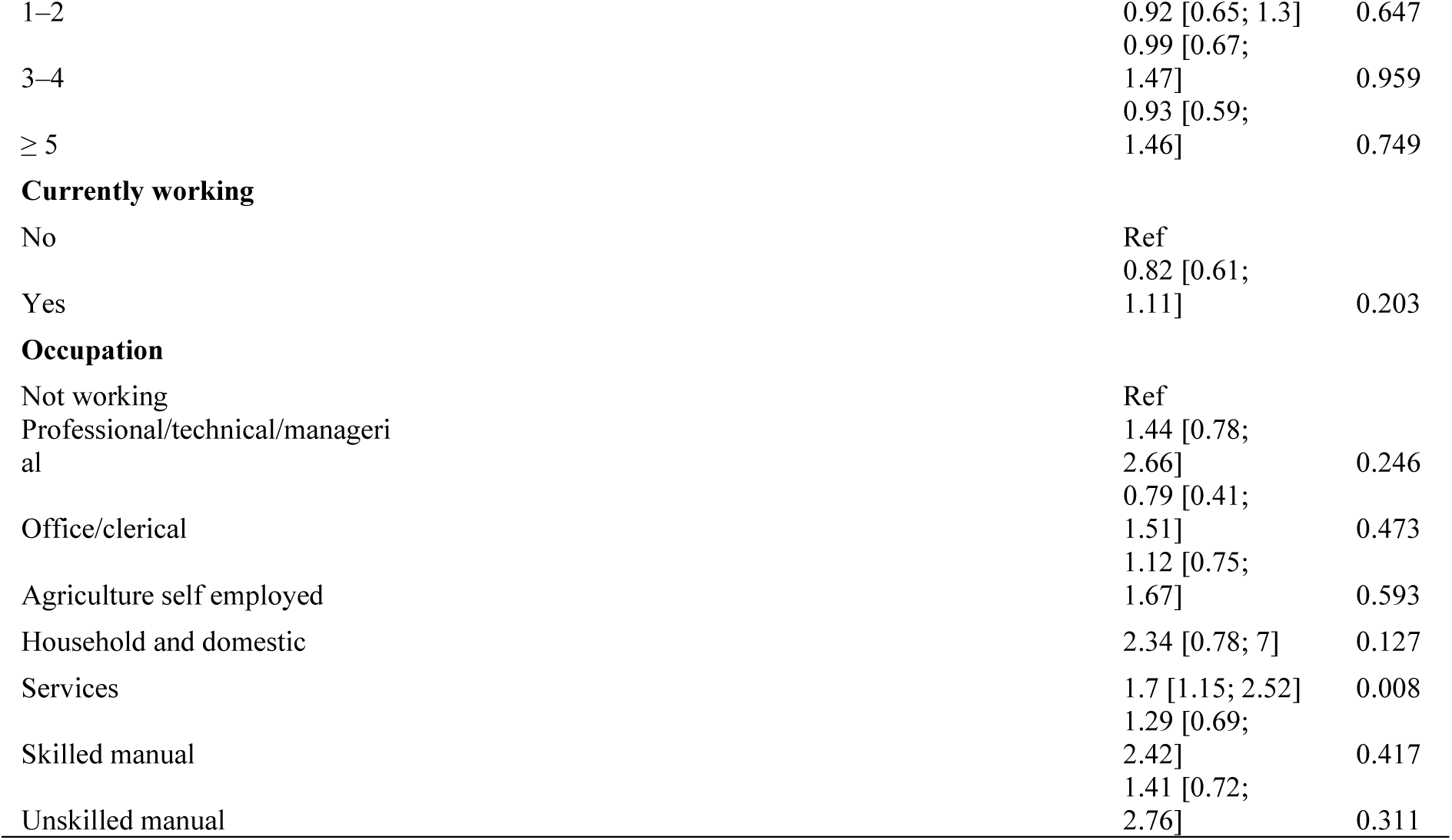
Multivariate result from the block wise models I, II and III (complementary to table 3)

**Annex 8 :** Fit indices for successive multivariate models based on conceptual blocks.

| Model | McFadden R<sup>2</sup> | Deviance | Null Deviance | Delta R<sup>2</sup> |
| --- | --- | --- | --- | --- |
| Model I (Context) | 0.1534 | 5185.78362 | 6125.38485 |  |
| Model II (Household) | 0.155 | 5173.02536 | 6121.64174 | 0.0016 |
| Model III (Individual characteristics) | 0.172 | 4979.57976 | 6014.34141 | 0.017 |
| Model IV (Nutritional and health behavioural) | 0.1758 | 4956.78734 | 6014.34141 | 0.0038 |

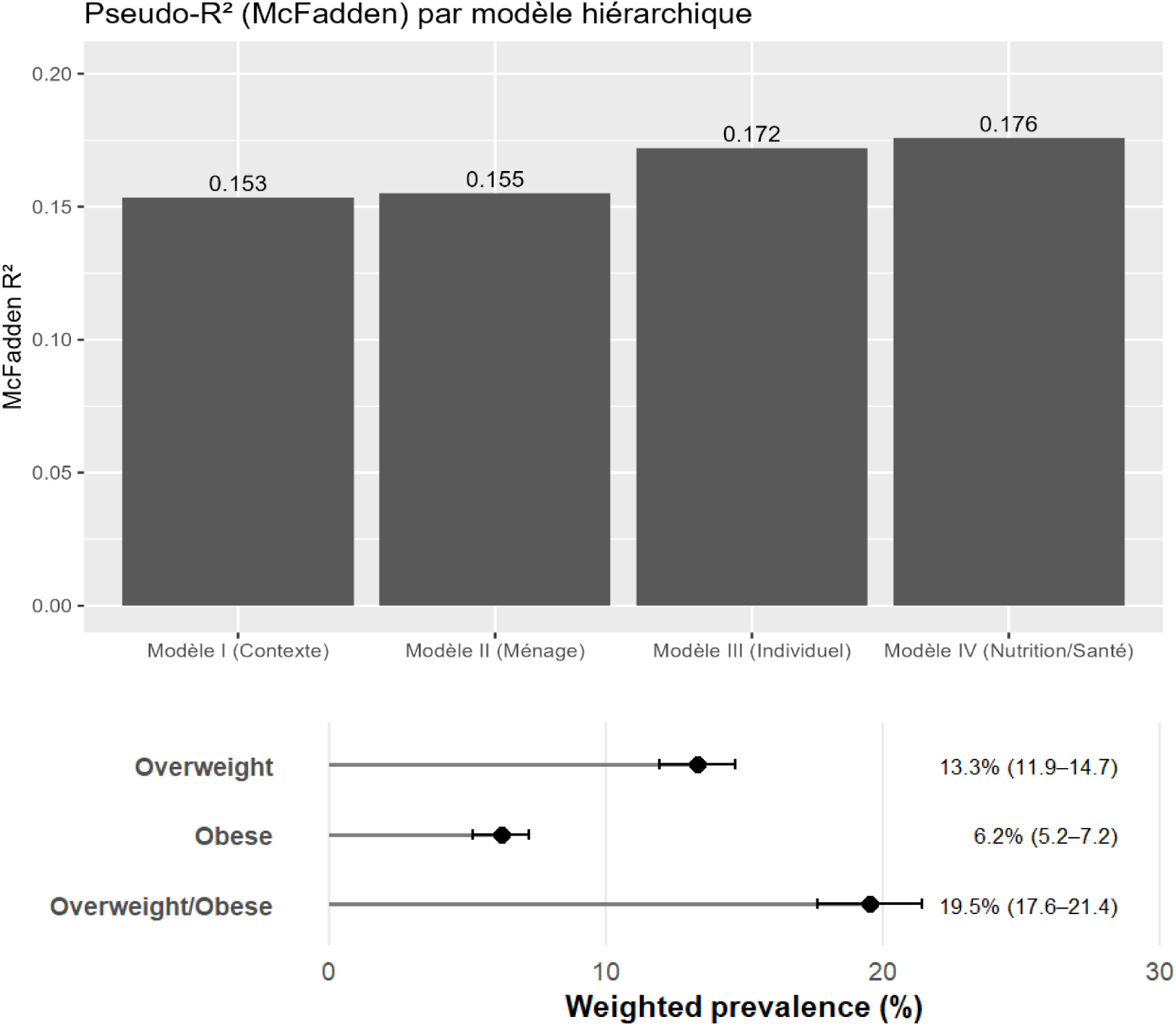

**Annex 9 :** Unweighted and weighted prevalence of overweight and obesity among women of reproductive age.

| N=10740 | n | Unweight (%) | Weighted (%) | 95%CI low | 95%CI High |
| --- | --- | --- | --- | --- | --- |
| Overweight | 1136 | 10.57 | 13.31 | 11.95 | 14.67 |
| Obese | 439 | 4.08 | 6.22 | 5.22 | 7.21 |
| Overweight/obese | 1575 | 14.66 | 19.53 | 17.64 | 21.42 |

**Annex 10 :** Unweighted and weighted prevalence of overweight/obesity by province among women of reproductive age.

| Province | Cases (n) | Total (N) | Prevalence (%) | Weighted (%) | 95% CI lower | 95% CI upper |
| --- | --- | --- | --- | --- | --- | --- |
| Kinshasa | 247 | 619 | 39.9 | 38 | 32.8 | 43.2 |
| Nord-Kivu | 182 | 563 | 32.3 | 33 | 27.8 | 38.3 |
| Haut-Katanga | 128 | 479 | 26.7 | 28.9 | 20.3 | 37.6 |
| Sud-Kivu | 105 | 537 | 19.6 | 22.8 | 9.12 | 36.4 |
| Haut-Uele | 62 | 255 | 24.3 | 21.9 | 14.4 | 29.3 |
| Bas-Uele | 50 | 282 | 17.7 | 19.8 | 15.2 | 24.4 |
| Tshopo | 52 | 362 | 14.4 | 18.1 | 9.99 | 26.2 |
| Maniema | 58 | 379 | 15.3 | 17.7 | 10.6 | 24.8 |
| Equateur | 60 | 405 | 14.8 | 16.8 | 8.97 | 24.7 |
| Lualaba | 43 | 372 | 11.6 | 16.4 | 10.1 | 22.7 |
| Ituri | 84 | 542 | 15.5 | 14.3 | 8.58 | 20 |
| Kasaï Oriental | 47 | 391 | 12 | 14.1 | 9.04 | 19.1 |
| Nord-Ubangi | 42 | 362 | 11.6 | 12.7 | 6.7 | 18.7 |
| Mongala | 53 | 365 | 14.5 | 11.5 | 7.13 | 15.8 |
| Kwilu | 33 | 454 | 7.27 | 10.6 | 5.16 | 16.1 |
| Mai-Ndombe | 36 | 337 | 10.7 | 9.9 | 5.72 | 14.1 |
| Lomami | 29 | 390 | 7.44 | 9.5 | 3.98 | 15 |
| Kasaï Central | 37 | 477 | 7.76 | 9.49 | 4.03 | 15 |
| Kongo Central | 54 | 441 | 12.2 | 9.3 | 5.36 | 13.3 |
| Kasaï | 24 | 446 | 5.38 | 9.27 | 0 | 18.8 |
| Sud-Ubangi | 46 | 517 | 8.9 | 8.9 | 3.01 | 14.8 |
| Tanganyika | 27 | 406 | 6.65 | 7.05 | 2.33 | 11.8 |
| Haut-Lomami | 33 | 472 | 6.99 | 6.82 | 4.23 | 9.41 |
| Sankuru | 19 | 286 | 6.64 | 5.75 | 2.66 | 8.84 |
| Kwango | 14 | 325 | 4.31 | 3.95 | 0.25 | 7.65 |
| Tshuapa | 10 | 276 | 3.62 | 2.21 | 0.08 | 4.34 |

**Annex 11 :** Sensitivity analysis (Stability of AoR Model IV comparing women 15–49-year-olds versus Women 20-49 years)

| Covariate | aOR (15–49 years)<br>[95% CI] | p-value<br>(15–49) | aOR (20–49 years)<br>[95% CI] | p-value<br>(20–49) | %<br>change | Stable<br>(<20%) |
| --- | --- | --- | --- | --- | --- | --- |
| Wealth: Richest (ref Poorest) | 4.21 [2.45–7.22] | 2.27E-07 | 4.83 [2.66–8.77] | 2.96E-07 | 12.8 | TRUE |
| Education: Higher (ref None) | 1.29 [0.77–2.16] | 0.331 | 1.20 [0.68–2.09] | 0.53 | 7.9 | TRUE |
| Residence: Rural (ref Urban) | 0.61 [0.48–0.79] | 1.76E-04 | 0.57 [0.44–0.74] | 3.69E-05 | 7.7 | TRUE |
| Education: Secondary (ref None) | 1.38 [0.98–1.96] | 0.0672 | 1.30 [0.89–1.90] | 0.173 | 6.5 | TRUE |
| Wealth: Rich (ref Poorest) | 2.41 [1.62–3.60] | 1.78E-05 | 2.57 [1.71–3.87] | 6.11E-06 | 6.2 | TRUE |
| Any unhealthy food consumed | 0.97 [0.80–1.17] | 0.74 | 1.01 [0.83–1.24] | 0.906 | 4.4 | TRUE |
| Wealth: Poor (ref Poorest) | 1.26 [0.86–1.86] | 0.231 | 1.32 [0.89–1.96] | 0.166 | 4.3 | TRUE |
| MDD-W met ($\geq 5$ food groups) | 1.22 [0.95–1.56] | 0.112 | 1.17 [0.92–1.50] | 0.205 | 4.3 | TRUE |
| Household electricity: Yes (ref No) | 1.31 [0.97–1.76] | 0.0781 | 1.28 [0.95–1.74] | 0.106 | 1.8 | TRUE |
| Wealth: Middle (ref Poorest) | 1.65 [1.13–2.41] | 0.00955 | 1.64 [1.10–2.46] | 0.0155 | 0.4 | TRUE |

**Annex 12:**
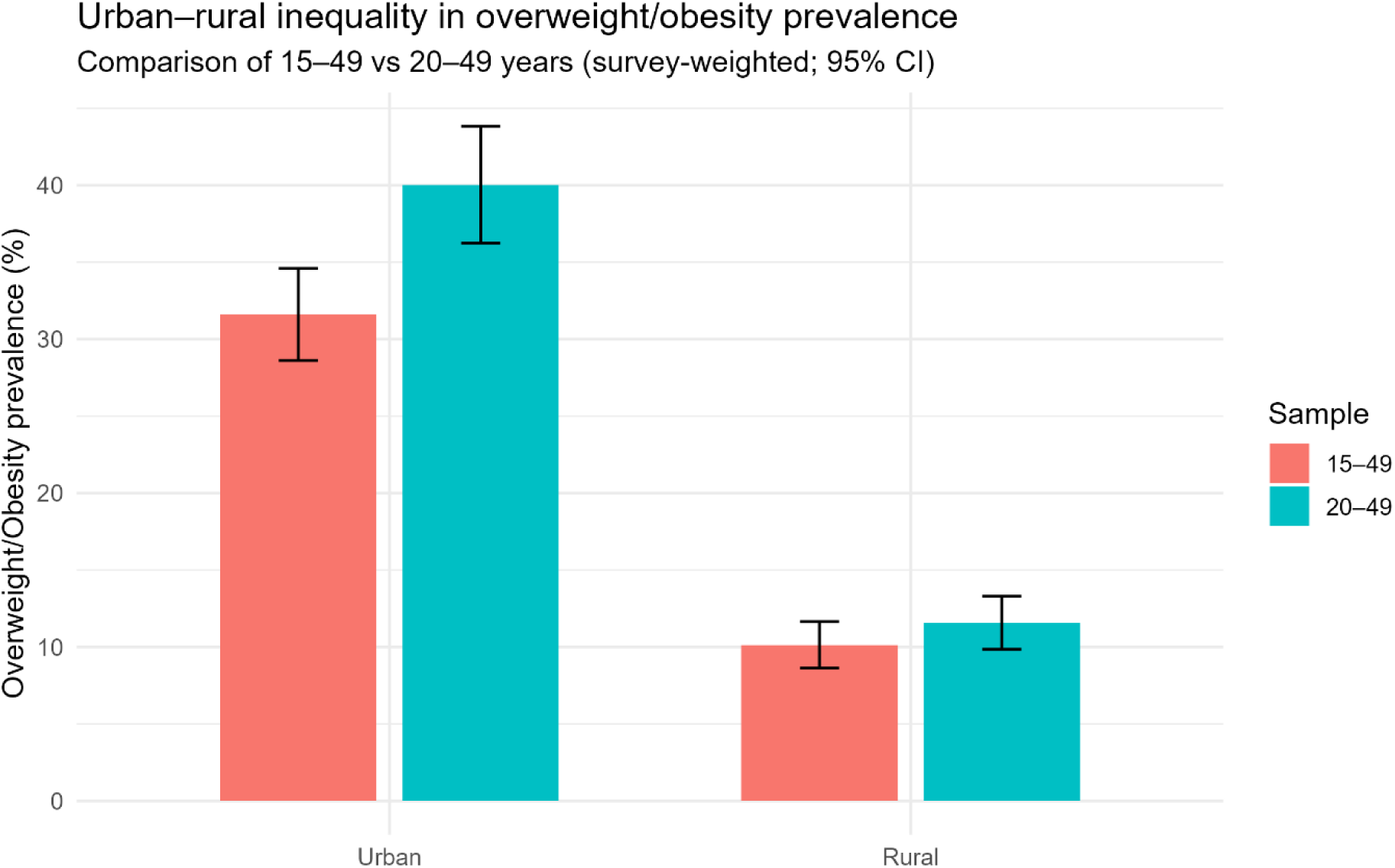
Prevalence of overweight/obesity by age group and residence area.

## Références

1. Ng M, Gakidou E, Lo J, Abate YH, Abbafati C, Abbas N, et al. Global, regional, and national prevalence of adult overweight and obesity, 1990–2021, with forecasts to 2050: a forecasting study for the Global Burden of Disease Study 2021. Lancet. 2025 Mar 8;405(10481):813–38.

2. World Health Organization. Obesity and overweight. Geneva : WHO; 2023. Available from: https://www.who.int/news-room/fact-sheets/detail/obesity-and-overweight

3. Popkin BM, Corvalan C, Grummer-Strawn LM. Dynamics of the double burden of malnutrition and the changing nutrition reality. Lancet [Internet]. 2020 Jan 4 [cited 2025 May 18];395(10217):65–74. Available from: https://www.sciencedirect.com/science/article/abs/pii/S0140673619324973

4. Masters WA, Finaret AB, Block SA. The economics of malnutrition: Dietary transition and food system transformation. Handb Agric Econ. 2022 Feb 5 [cited 2025 Aug 5];6:4997–5083.

5. Popkin BM, Ng SW. The nutrition transition to a stage of high obesity and noncommunicable disease prevalence dominated by ultra-processed foods is not inevitable. Obes Rev. 2022;23(1):1–18.

6. Popkin BM, Adair LS, Ng SW. Global nutrition transition and the pandemic of obesity in developing countries. Nutr Rev. 2012 Jan;70(1):3–21.

7. Davis JN, Oaks BM, Engle-Stone R. The Double Burden of Malnutrition: A Systematic Review of Operational Definitions. Curr Dev Nutr [Internet]. 2020 Sep 1 [cited 2025 Aug 5];4(9):nzaa127. Available from: https://pmc.ncbi.nlm.nih.gov/articles/PMC7456307/

8. World Health Organization. The double burden of malnutrition: policy brief. WHO. Geneva; 2017.

9. Wells JC, Sawaya AL, Wibaek R, Mwangome M, Poullas MS, Yajnik CS, et al. The double burden of malnutrition: aetiological pathways and consequences for health. Lancet. 2020;395(10217):75–88.

10. Alem AZ, Yeshaw Y, Liyew AM, Tessema ZT, Worku MG, Tesema GA, et al. Double burden of malnutrition and its associated factors among women in low and middle income countries: findings from 52 nationally representative data. BMC Public Health. 2023;23(1):1–16. Available from: 10.1186/s12889-023-16045-4

11. Ozodiegwu ID, Littleton MA, Nwabueze C, Famojuro O, Quinn M, Wallace R, et al. A qualitative research synthesis of contextual factors contributing to female overweight and obesity over the life course in sub-Saharan Africa. PLoS One. 2019 Nov 4;14(11): e0224612. Doi: 10.1371/journal.pone.0224612

12. Christian AK, Dake FAA. Profiling household double and triple burden of malnutrition in sub-Saharan Africa: prevalence and influencing household factors. Public Health Nutr. 2022;25(6):1563–76.

13. Tamir TT, Mekonen EG, Workneh BS, Techane MA, Terefe B, Zegeye AF. Overnutrition and associated factors among women of reproductive age in Sub-Saharan Africa: A hierarchical analysis of 2019–2023 standard demographic and health survey data. Nutrition [Internet]. 2024 Dec 1 [cited 2025 Aug 2];128:112563.

14. Atsu P, Mohammed A, Adu C, Aboagye RG, Ahinkorah BO, Seidu AA. Residence-based inequalities in overweight/obesity in sub-Saharan Africa: a multivariate non-linear decomposition analysis. Trop Med Health. 2024; 52:13. doi:10.1186/s41182-024-00593-5

15. Kandala NB, Stranges S. Geographic variation of overweight and obesity among women in Nigeria: A case for nutritional transition in Sub-Saharan Africa. PLoS One. 2014 Jun 30;9(6).

16. Amugsi DA, Dimbuene ZT, Mberu B, Muthuri S, Ezeh AC. Prevalence and time trends in overweight and obesity among urban women : an analysis of demographic and health surveys data from 24 African countries, 1991 – 2014. BMJ Open. 2017 Oct 27;7(10): e017344. doi: 10.1136/bmjopen-2017-017344. PMID: 29079606; PMCID: PMC5665233.

17. Beyene ET, Cha S, Jin Y. Overweight and obesity trends and associated factors among reproductive women in Ethiopia. Glob Health Action [Internet]. 2024 [cited 2025 Dec 15];17(1):2362728. Available from: https://pmc.ncbi.nlm.nih.gov/articles/PMC11172244/

18. Adam J, Luoga P, Mtawa A, Nyamhanga T. Prevalence and associated factors of overweight and obesity among adult women in Tanzania from the 2022 Tanzania Demographic and Health Survey. Sci Reports 2025 151 [Internet]. 2025 Sep 30 [cited 2025 Dec 15];15(1):33978-. Available from: https://www.nature.com/articles/s41598-025-11141-4

19. Asosega KA, Aidoo EN, Adebanji AO, Owusu-Dabo E. Examining the risk factors for overweight and obesity among women in Ghana: A multilevel perspective. Heliyon [Internet]. 2023 May 1 [cited 2025 Dec 15];9(5):e16207. Available from: https://www.sciencedirect.com/science/article/pii/S240584402303414X

20. Jaacks LM, Vandevijvere S, Pan A, McGowan CJ, Wallace C, Imamura F, et al. The obesity transition: stages of the global epidemic. Lancet Diabetes Endocrinol [Internet]. 2019 Mar 1 [cited 2025 Dec 14];7(3):231–40. Available from: https://www.thelancet.com/action/showFullText?pii=S2213858719300269

21. Ministère de la Santé Publique Hygiene et Prévention. Enquête Démographique et de Santé en République Démocratique du Congo 2013-2014. Kinshasa (Republique Democratique du Congo) et Rockville, Maryland (USA) : Ministère de la santé publique, hygiene et prevention, MSPHP et ICF International; 2014.

22. Institut national de la statistique et École de santé publique de Kinshasa. Rapport de l’enquête démographique et de santé EDS-RDC III 2023–2024. Kinshasa, République Démocratique du Congo; 2025.

23. Makulo JR, Tshiswaka Mutombo T, Kamitalu Kabongo R, Tunda AK, Sumaili EK, Kisoka Lusunsi C, et al. Double Burden of Malnutrition and Validity of Overweight/Obesity Indicators Among Students in the Democratic Republic of Congo: A Cross Sectional Study From the University of Kinshasa. Diabetes Metab Syndr Obes. 2025;18(June):2057–65.

24. Wakilongo W, Abbeddou S, Vanhoutte L, Amougou N, Mubagwa M, Elmira C, et al. Biocultural determinants of overweight-obesity among adult women experiencing the nutritional transition in the Democratic Republic of Congo. Front Nutr. 2024;11.

25. Hawkes C, Ruel MT, Salm L, Sinclair B, Branca F. Double-duty actions: seizing programme and policy opportunities to address malnutrition in all its forms. Lancet [Internet]. 2020 Jan 11 [cited 2025 Sep 5];395(10218):142–55. Available from: https://pubmed.ncbi.nlm.nih.gov/31852603/

26. Mcleroy KR, Bibeau D, Steckler A, Glanz K. An ecological perspective on health promotion programs. Health Educ Q [Internet]. 1988 [cited 2025 Dec 16];15(4):351–77. Available from: https://pubmed.ncbi.nlm.nih.gov/3068205/

27. Pradeilles R, Holdsworth M, Olaitan O, Irache A, Osei-Kwasi HA, Ngandu CB, et al. Body size preferences for women and adolescent girls living in Africa: a mixed-methods systematic review. Public Health Nutr. 2022;25(3):738–59.

28. Van der Pligt P, Willcox J, Hesketh KD, Ball K, Wilkinson S, Crawford D, et al. Systematic review of lifestyle interventions to limit postpartum weight retention: implications for future opportunities to prevent maternal overweight and obesity following childbirth. Obes Rev [Internet]. 2013 Oct [cited 2025 Dec 16];14(10):792–805. Available from: https://pubmed.ncbi.nlm.nih.gov/23773448/

29. World health organization. WHO growth reference data for 5–19 years: BMI-for-age (z-scores) [Internet]. WHO. 2007 [cited 2025 Dec 21]. Available from: https://www.who.int/tools/growth-reference-data-for-5to19-years/indicators/bmi-for-age

30. Ramalan MA, Gezawa ID, Uloko AE. Prevalence and trends of adult overweight and obesity in Nigeria : a systematic review and meta-analysis. Niger J Clin Pract. 2019;22(8):1070–7.

31. World Health Organisation-Best buys. “Best buys” and other recommended interventions for the prevention and control of noncommunicable diseases. Geneva : WHO;. 2017;17(9):28. Available from: https://iris.who.int/handle/10665/259232.

32. Willett WC. Nutritional epidemiology. 3rd ed. (Monographs in Epidemiology and Biostatistics,; vol 40). New York: Oxford University Press; 2012. 552p.

33. Gibson RS. Principles of Nutritional Assessment [Internet]. 2nd ed. Oxford (UK): oxford University Press ; 2005 [cited 2025 Dec 16]. 928 p.

